# A reproducibility evaluation of the effects of MRI defacing on brain segmentation

**DOI:** 10.1101/2023.05.15.23289995

**Authors:** Chenyu Gao, Bennett A. Landman, Jerry L. Prince, Aaron Carass

## Abstract

**Purpose:** Recent advances in magnetic resonance (MR) scanner quality and the rapidly improving nature of facial recognition software have necessitated the introduction of MR defacing algorithms to protect patient privacy. As a result, there are a number of MR defacing algorithms available to the neuroimaging community, with several appearing in just the last five years. While some qualities of these defacing algorithms, such as patient identifiability, have been explored in previous works, the potential impact of defacing on neuroimage processing has yet to be explored.

**Approach:** We qualitatively evaluate eight MR defacing algorithms on 179 subjects from the OASIS-3 cohort and the 21 subjects from the Kirby-21 dataset. We also evaluate the effects of defacing on two neuroimaging pipelines— SLANT and FreeSurfer—by comparing the segmentation consistency between the original and defaced images.

**Results:** Defacing can alter brain segmentation and even lead to catastrophic failures, which are more frequent with some algorithms such as *Quickshear*, *MRI_Deface*, and *FSL_deface*. Compared to FreeSurfer, SLANT is less affected by defacing. On outputs that pass the quality check, the effects of defacing are less pronounced than those of rescanning, as measured by the Dice similarity coefficient.

**Conclusions:** The effects of defacing are noticeable and should not be disregarded. Extra attention, in particular, should be paid to the possibility of catastrophic failures. It is crucial to adopt a robust defacing algorithm and perform a thorough quality check before releasing defaced datasets. To improve the reliability of analysis in scenarios involving defaced MRIs, it’s encouraged to include multiple brain segmentation pipelines.

## 1 Introduction

Magnetic resonance (MR) images are widely used to study the brain and there has been an ever increasing number of whole head MRIs being acquired clinically—about 40 million^1^ scans annually in the United States.^1^ In conjunction with the increasing number of scans are three important trends. First, there have been considerable improvements in scanner technology including resolution and signal-to-noise ratio (SNR) improvements which have come from many factors including the increased proportion of 3 Tesla (3T) scanners over 1.5T systems and the use of compressed sensing image acquisition. These improvements have led to increasing numbers of clinical scans that are reconstructed with high fidelity. Second, there has been increasing efforts by medical imaging stake holders towards open and reproducible science,^2^ which has led to ever increasing amounts of acquired whole-head MRIs being made publicly available. These open data initiatives are aimed at reducing barriers to entry in many research fields requiring medical images of the human brain. Studies like ABIDE,^3^ ADNI,^4^ and HCP^5^ are all examples of large studies that have made a considerable amount of their data publicly available, and there are many others.^6–10^ Additionally, many medical health systems are commoditizing patient data by deidentifying it and selling the data to commercial entities.^11, 12^ Third, deep learning (DL) technologies, fueled by vast training data, architectural advancements, and remarkable computational power improvements, have paved the way for more sophisticated face recognition capabilities. In this context, issues surrounding privacy have become prominent.

Investigators are provided guidelines by their institutional review boards (IRBs) for handling protected health information (PHI) that is collected during a human subjects research study. PHI comprising textual information (e.g., meta-data such as name, date of birth, medical record number, etc.), for example, can be readily removed while exporting data from a Picture Archiving and Communication System (PACS) or after export using one of several software packages.^13, 14^ The question of whether and how investigators should handle the facial information that is present in high quality medical images is not yet agreed upon. In particular, although some studies have shown that photographs can be matched with reconstructions from high-quality medical images;^15–17^ these matches are made in highly-controlled, small-scale settings, in which there is always a corresponding pair between photograph and MR reconstruction. The capability to match photographs with reconstructions from MRIs is not routinely available, nor has it been tested on large scale cohorts where matches may not exist. Nevertheless, it is reasonable to assume that with technological advances, this capability could extend to collections of photographs found on the internet.

Many algorithms have been developed over the past 15 years to tackle privacy issues associated with facial information in medical images.^18–26^ These algorithms operate by obscuring or removing potentially recognizable portions of the face from the MR images, thus reducing the utility of 3D reconstruction for identification purposes. We refer to them collectively as “defacing algorithms” although some also remove the ears as well. While defacing algorithms remove facial features to preserve privacy, concerns have arisen that they may negatively affect analysis. Sitter et al. showed that automated pipelines for volumetric analysis exhibit a higher failure rate when applied to defaced images as compared to original images.^27^ Moreover, Buimer et al. found that the effects of defacing vary across the subject’s age and across brain regions.^28^ Other studies have also shown the effects of defacing on downstream tasks, such as co-registration between MR images and EEG/MEG data,^29^ brain atrophy estimation,^30^ quality measurements,^31^ whole brain segmentation,^32^ volume analysis,^25^ and head and neck cancer segmentation.^33^ Theyers et al. conducted a comprehensive study on the effects of defacing on brain volume measurements and fMRI preprocessing, as well as image registration across multiple cohorts.^34^ They found that, beyond the direct errors caused by defacing, none of the resulting differences were significantly greater than those that could be introduced by using different DICOM-to-NIfTI converters.

The findings from these studies help us to better understand the consequences of applying defacing techniques and to guide us towards privacy protection standards that the entire community can agree upon. But further characterization is necessary. To our knowledge, no prior studies have analyzed the effects of defacing on brain segmentation using multiple advanced pipelines, each with a fundamentally different methodological approach. Moreover, few studies have simultaneously i) used a substantial volume of data for validation and ii) included a comprehensive selection of defacing algorithms for comparison.

In this work, we include eight defacing algorithms that cover the majority of publicly available choices for defacing (See Fig. 1 for examples), and use MR images from 200 subjects across two public datasets. As part of our evaluation, we analyze the effects these defacing algorithms have on the performance of two popular neuroimaging pipelines: SLANT^35, 36^ and FreeSurfer.^37^ In our experiments, we begin by applying each defacing algorithm to T_1_-weighted MR images of 179 subjects from the OASIS-3 cohort.^8^ We conduct a manual quality check on the outputs of each defacing algorithm, in which we identify success and two types of failures. We exclude those failure cases from further analysis, as described in Sec. 3.1. Next, we feed both the defaced images and the original images into two brain segmentation pipelines (SLANT and FreeSurfer). To measure the effects of defacing, we compute the Dice Similarity Coefficient (DSC)^38^ between the segmentations obtained from the original MR image and the corresponding defaced MR image. To quantify our original *vs.* defaced segmentation results, we compare the effects of defacing with those of the segmentation of scan-rescan data—after scan-rescan subject alignment—by using the Kirby-21 dataset.^39^

**Fig 1:**
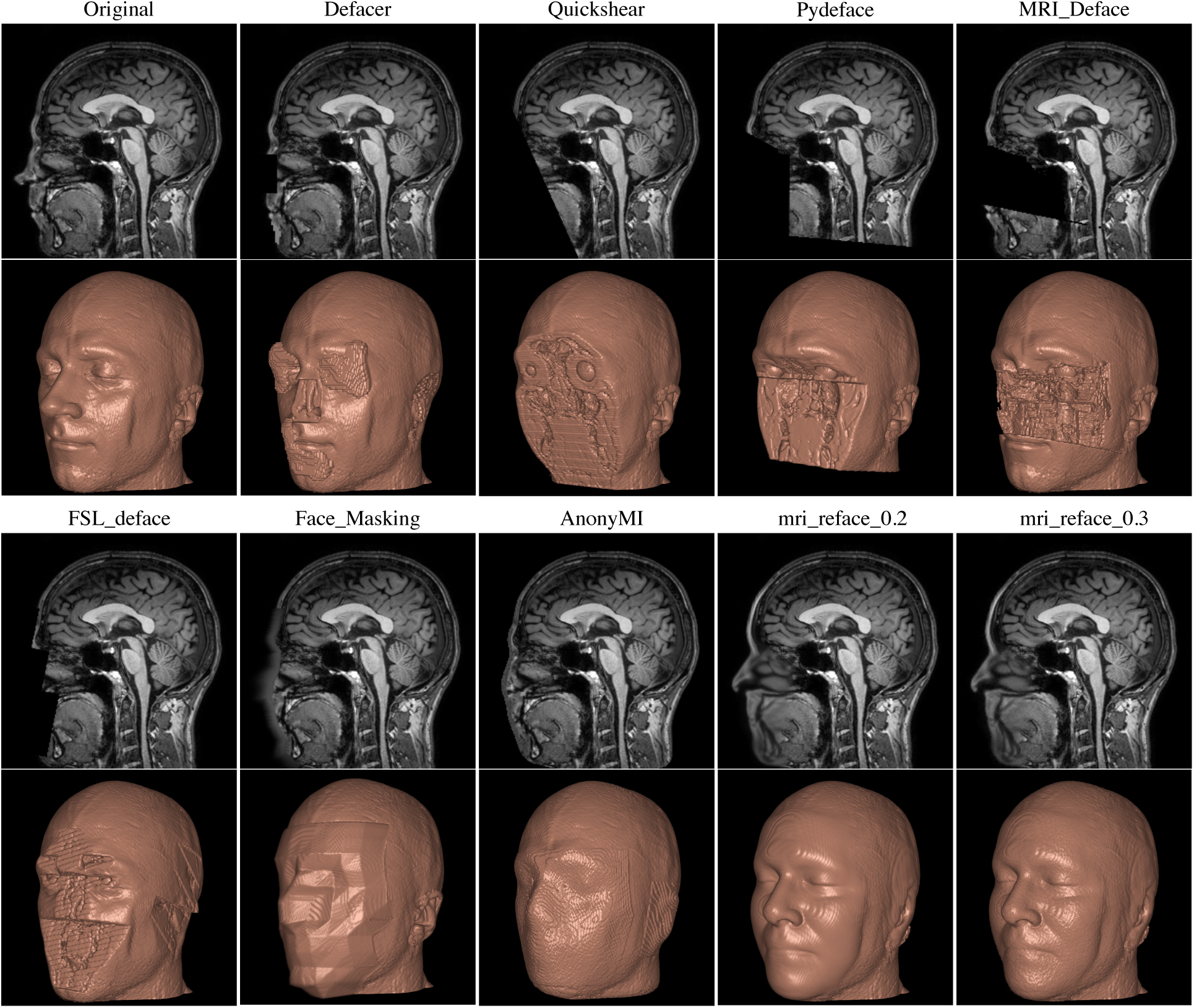
A sagittal slice of an MRI is displayed over the reconstruction of the whole head MRI. Top set of images from left to right is: the acquired MRI and then defacing using *Defacer*,^23^ *Quick-Shear*,^19^ *Pydeface*,^22^ and *MRI_Deface*.^18^ The bottom set of images from left to right is: defacing using *FSL_Deface*,^21^ *Face_Masking*,^20^ *AnonyMI*,^24^ *mri_reface_0.2*,^25^ and *mri_reface_0.3*.^25^

We observe that defacing has a measurable impact on brain segmentation, with the effects being larger on FreeSurfer segmentations (DSC = 0.918 *±* 0.019) than on SLANT segmentation (DSC = 0.970*±*0.005). Also, we found that DSC = 0.879*±*0.015 for FreeSurfer segmentation on scan–rescan pairs and DSC = 0.952 *±* 0.005 for SLANT segmentation on scan-rescan pairs. From this result we conclude that the effects of defacing are smaller than those of scan–rescan followed by registration. Based on this, one might be tempted to ignore the effects of defacing. But we also found that catastrophic failures in brain segmentation can be caused by defacing. While most of these failures are typically easy to detect during quality checks, some can have subtle effects, as shown in Sec. 5.3 and discussed in Sec. 5.4.

## 2 Methods and Materials

### 2.1 Defacing Algorithms

The eight defacing algorithms used in our comparison are described in detail in the Supplementary Material. Here, we provide a brief outline of the eight methods.

*Defacer*^23^ is an open-source deep-learning based method for MRI anonymization. It uses a deep network to identify the eyes, ears, nose, and mouth in an MR image. This is followed by image processing techniques to manipulate the intensity values of the detected facial feature voxels and their immediate surroundings.

*Quickshear*^19^ uses a precomputed brain mask and edge detection to identify a “shearing plane” that separates the face from the brain. We generate the brain masks required by *Quickshear* using BET (Brain Extraction Tool)^40^ with default settings.

*MRI_Deface*,^18^ *Pydeface*,^22^ and *FSL_deface*^21^ have a similar workflow, in that they each register a template with a corresponding mask (or masks) to the input image; with non-brain voxels being masked out or manipulated through some straightforward image processing to provide anonymization. The methods use linear registration with either Fischl et al.^41^ or FLIRT.^42^ *FSL_deface* has a key difference with *MRI_Deface* and *Pydeface* in that it includes the ears in its defacing mask. Given the identifiability of the ears,^43^ this seems like an unfortunate oversight of *MRI_Deface* and *Pydeface*.

*Face_Masking*^20^ focuses on blurring the facial surface so as not to introduce hard intensity edges to the whole head MRI that can confuse subsequent processing tools. The result is an artificial *cubist*-like face, see Fig. 1 for an example.

*AnonyMI*^24^ uses a combination of a watershed algorithm^44^ and a non-linear registration^45^ of a template, to identify the facial surface and features. The template provides a generic face, while the facial surface and features are blended with the generic face while ensuring that the image intensities in the blended regions come from the same distribution as the facial features.

*mri_reface*^25^ uses a non-linear registration^46^ to bring an average face template into the space of the input image and then replaces the facial features with those of the template. The average face template intensities are transformed to match the input intensities with intensity matching—similar to the one described by Nyúl and Udupa^47^—followed by bias correction for smooth local intensity normalization between the images.^48^ We use both publicly available versions of *mri_reface* in the present study.

We note that Schwarz et al. evaluated whether MR images were correctly recognizable by facial recognition software after defacing.^25^ Of the presented methods in that paper, the ranking in terms of correct matching between photos and MRIs of participants after defacing was: *FSL_deface* (28%); *mri_reface* (30%); *MRI_Deface* (33%); *PyDeface* (38%). The authors also included an intra-class correlation coefficient (ICC) comparison between the presented methods before and after defacing. ICC would identify brain structures that have changed their volume in some way, it would not highlight changes in the spatial positioning of those brain structures which is critically important when potentially considering the accidental removal of portions of the brain due to defacing. We note this, as we have observed brain structures “*moving*” if their segmentation is performed on the original or defaced images, see Fig. 6 for an example.

### 2.2 Dataset

All the defacing methods in our study have reported results on T_1_-w MRIs; additionally T_1_-w MRIs are typically the images acquired at the highest resolution—important for correct facial recognition—and are also the most commonly acquired images. As such, we have focused our defacing evaluation on T_1_-w MRIs from a subset of the OASIS-3^8^ cohort and the complete set of the Kirby-21^39^ study. All images are reoriented to match the approximate orientation of the standard template images (MNI152) using `fslreorient2std`,^49^ which only applies 0, 90, 180 or 270 degree rotations.

#### 2.2.1 OASIS-3^8^

OASIS-3 is the third release of the Open Access Series of Imaging Studies (OASIS)^50^ and includes retrospective data from 1,379 individuals collected over a period of 30 years by the WUSTL Knight ADRC. Of these 1,379 individuals, 755 were cognitively normal adults, while the remaining individuals were at different stages of cognitive decline and ranged in age from 42 to 95 years. OASIS-3 comprises over 2,800 imaging sessions that include T_1_-w, T_2_-w, FLAIR, ASL, SWI, resting-state BOLD, and DTI. For our study, we selected a random sample of 179 subjects from the OASIS-3 cohort. For each of these subjects, we randomly selected one T_1_-w image from the available imaging sessions to use in our experiments. See the Supplemental Material for the list of selected images.

#### 2.2.2 Kirby-21^39^

Kirby-21 is part of the Multi-Modal MRI Reproducibility Resource and includes scan-rescan imaging sessions of 21 healthy subjects with no history of neurological conditions. Each subject underwent two identical 1-hour scanning sessions, with a short break in between and repositioning before the second session. The resulting dataset comprises 42 sessions that include MPRAGE, FLAIR, DTI, resting state fMRI, and so on. For our study, we used the MPRAGE images from the scan-rescan sessions of the 21 subjects.

### 2.3 Brain Segmentation Pipelines

We include two popular whole head MRI segmentation pipelines to evaluate the effects of defacing on brain segmentation.

#### 2.3.1 SLANT^35, 36^

The spatially localized atlas network tiles (SLANT) method employs multiple independent 3D convolutional networks for segmenting the brain. Each of the networks is only responsible for a particular spatial region, thus the task of each network is simplified to focus on patches from a similar portion of the brain. To enable this, affine registration, N4 bias field correction, and intensity normalization are employed to roughly normalize each brain to the same space before segmentation. After each network performs its duty, the segmentation labels are fused together to form the final labels for the 132 anatomical regions of the brain. SLANT is publicly^2^ available and is reported to have high intra- and inter-scan protocol reproducibility.^51^ In this study, we use version `1.0.3` with GPU support.

#### 2.3.2 FreeSurfer^37^

FreeSurfer is a widely used tool in the neuroimaging community that provides automated processing of MRI data to obtain measurements of various brain structures. In this study, we use the segmentation file, `aparc+aseg.mgz`, generated by FreeSurfer’s “`recon-all`” method. The file contains information about the cortical regions and subcortical structures segmented by FreeSurfer, including the Desikan-Killiany atlas-based parcellation of the cerebral cortex and the segmentation of subcortical structures such as left and right caudate, putamen, pallidum, thalamus, lateral ventricles, hippocampus, and amygdala. We use version `7.3.2`.

## 3 Experimental Setup

### 3.1 Defacing Quality Check

In Sec. 4, we present the results of application of the eight defacing algorithms on our total of 200 images (179 T_1_-weighted images from OASIS-3 and 21 MPRAGE images from the first session of each subject in the Kirby-21). We initially review the resulting defaced images manually; this review is a first stage quality check that is focused on identifying any problems in the images. Each image is checked by viewing the axial slices from lateral to superior and then again from superior to lateral. This initial review classifies the output of all eight defacing algorithms into three categories:

#### Success

The particular defacing algorithm processes the MR image as expected. Although some facial voxels that are supposed to be removed may remain, there are not any unrecoverable errors as observed in the other two categories.

#### Type I Failure

The defacing algorithm fails to detect facial features or run properly. As a consequence, the face remains untouched and may still be recognizable.

#### Type II Failure

Some non-zero proportion of the brain is removed due to the excesses of the defacing algorithm.

### 3.2 Quantify the Effects of Defacing

To quantify the effects of the defacing algorithms, we first applied them—using their default parameters—to the 179 T_1_-weighted MR images from OASIS-3. After the completion of the quality check outlined in Sec. 3.1, we include only those defacing results that are classified as “Success” for our subsequent analyses. We then run both SLANT and FreeSurfer on the original data as well as on the non-excluded output of each defacing algorithm. This results in segmentation label images from both SLANT and FreeSurfer for each of the subjects from OASIS-3 that passed our defacing quality check. Hereafter, we refer to the segmentation from the original data as the “*unaltered result*”, and the segmentation from the defaced data as the “*defacing result*”. We consider the segmentation result on the unaltered image as the ground truth, and compare the defacing result to this ground truth by calculating the Dice similarity coefficient (DSC),^38, 52^ using,

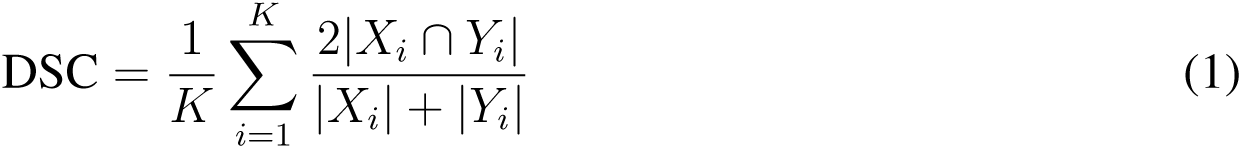

where *K* is the total number of labeled regions in the brain, *X_i_* and *Y_i_* are the binary segmentation masks for region *i* in the unaltered and defacing results, respectively. Here, *| · |* denotes the cardinality of the corresponding mask (i.e., the number of voxels).

### 3.3 Compare Defacing with Scan-Rescan

After quantifying the effects of defacing on segmentation by computing the DSC, we have a question:

#### Are these effects comparable or worse than those of rescanning a subject followed by registration?

To answer this question, we used the MPRAGE images from the scan-rescan sessions of the 21 subjects from the Kirby-21 cohort. This question is aimed at determining if any of the defacing methods have no more negative effect on SLANT and FreeSurfer than the variance associated with rescanning.

Firstly, we apply the eight defacing algorithms to the MPRAGE images from the first scan, using default parameters. We then exclude any defaced images that did not pass our quality check, see Sec. 3.1, by using only those images that are classified as “Success” for the subsequent steps. We then run SLANT and FreeSurfer, on both the original data and the defaced data. This enables us to obtain segmentation label images for each set of data. To quantify the effects of defacing on segmentation, we compute the DSC between the segmentation results obtained from the original and defaced data, as we did in the previous section. This is the same procedure that we used for the 179 OASIS-3 subject, as described in Sec. 3.2.

The availability of the second contemporaneous scan of each of the Kirby-21 participants allows us to take the additional step of registering the rescan (or second) MPRAGE image to the corresponding first MPRAGE of the same participant. We do this registration step using three different types of registration methods: rigid, affine, and deformable (SyN) implemented by ANTs.^46, 53^ After registration, we run SLANT and FreeSurfer on the rescan data registered to the first scan. We then compute the DSC to measure the difference between the segmentation results from the two scans.

## 4 Experimental Results

### 4.1 Quality Check

We report the results of our quality check in Table 1. *Quickshear* has a dramatic number of failure cases, most of which are “Type II Failure” cases—which is when some proportion of the brain is removed due to excessive (or inappropriate) defacing. We attribute this outcome to the dependence of *Quickshear* on the quality of the input brain mask. In our experiments, the brain masks were generated by BET without fine-tuning the parameters for each brain, and thus can be unreliable. Upon review, we regard the vast majority of *Quickshear* “Type II Failure” cases as a result of

**Table 1:** Manual Quality Check: The results of our manual quality check of all eight defacing algorithms across the 179 subjects of the OASIS-3 cohort^8^ and the 21 participants of the Kirby-21 dataset.^39^ Detailed results are included in the Supplemental Material.

| Algorithm | Dataset | Total | Success | Type I Failure | Type II Failure |
| --- | --- | --- | --- | --- | --- |
| *Defacer*<sup>23</sup> | OASIS-3 | 179 | 163 | 16 | 0 |
|  | Kirby-21 | 21 | 21 | 0 | 0 |
| *Quickshear*<sup>19</sup> | OASIS-3 | 179 | 78 | 0 | 101 |
|  | Kirby-21 | 21 | 20 | 0 | 1 |
| *Pydeface*<sup>22</sup> | OASIS-3 | 179 | 179 | 0 | 0 |
|  | Kirby-21 | 21 | 21 | 0 | 0 |
| *MRI_Deface*<sup>18</sup> | OASIS-3 | 179 | 136 | 7 | 36 |
|  | Kirby-21 | 21 | 21 | 0 | 0 |
| *FSL_deface*<sup>21</sup> | OASIS-3 | 179 | 142 | 0 | 37 |
|  | Kirby-21 | 21 | 21 | 0 | 0 |
| *Face_Masking*<sup>20</sup> | OASIS-3 | 179 | 176 | 0 | 3 |
|  | Kirby-21 | 21 | 21 | 0 | 0 |
| *AnonyMI*<sup>24</sup> | OASIS-3 | 179 | 179 | 0 | 0 |
|  | Kirby-21 | 21 | 21 | 0 | 0 |
| *mri_reface_0.2*<sup>25</sup> | OASIS-3 | 179 | 179 | 0 | 0 |
|  | Kirby-21 | 21 | 21 | 0 | 0 |
| *mri_reface_0.3*<sup>25</sup> | OASIS-3 | 179 | 179 | 0 | 0 |
|  | Kirby-21 | 21 | 21 | 0 | 0 |

BET and could possibly be rectified with an alternative skull-stripping software. However, we note that if the skull-stripping is done to a high enough standard then it can serve to deidentify an image and the additional step of defacing is superfluous. *Defacer* has the highest incidence of “Type I Failure” cases, wherein the algorithm encountered issues with detecting facial features, resulting in untouched faces in 16 subjects; *MRI_Deface* is the only other method to have “Type I Failure” cases, though it is only 7 of the 200 subjects we used in our studies—6 in OASIS-3 and 1 in Kirby-21. *Pydeface* has zero failure cases, unlike its counterparts—*FSL_deface* and *MRI_Deface*—that achieve defacing in a comparable manner, *i.e.*, by applying a predefined mask (or masks) after registration. *AnonyMI* and both versions of *mri_reface* also have zero failure cases. See the Supplemental Material for complete details about the quality check results.

### 4.2 OASIS-3

In our first experiment, we compare the performance of SLANT and FreeSurfer on unaltered images with their performance on defaced images. As outlined in Sec. 3.2, we applied the eight defacing algorithms to our OASIS-3 cohort of 179 subjects and then computed the DSC overlap that occurs between running SLANT (or FreeSurfer) on the unaltered images and the defaced images. In essence, we treat running SLANT (or FreeSurfer) on the unaltered images as a proxy for a gold standard segmentation; what underpins this approach is the assumption that defacing *does no harm* to the underlying brain data and *does not change* the position and orientation of the brain, in which case we can compare the segmentations of unaltered data to defaced data. In Fig. 2, we visualize these results as “raincloud” plots in which the dots are the mean DSC of the labels per subject, and the colors correspond to different defacing algorithms. The SLANT results have the “cloud” of the raincloud on the left, while the FreeSurfer results have the cloud on the right. We note that these results only include data that passed our quality check (see Sec. 3.1), which explains by *Quickshear* has considerably fewer “raindrops” than the other algorithms due to its large number of failure cases (see Table 1). We observe that for both SLANT and FreeSurfer across all tested defacing algorithms, there is not a single case where DSC = 1, which indicates that defacing always has an impact of the segmentation results from both SLANT and FreeSurfer. Also, the DSC from the SLANT segmentations are individually and collectively higher than those from the FreeSurfer segmentations, and the FreeSurfer segmentations have a considerably larger spread. Interestingly, neither segmentation algorithm depends strongly on which defacing method is used. A detailed discussion of these results is provided in Sec. 5.4.

**Fig 2:**
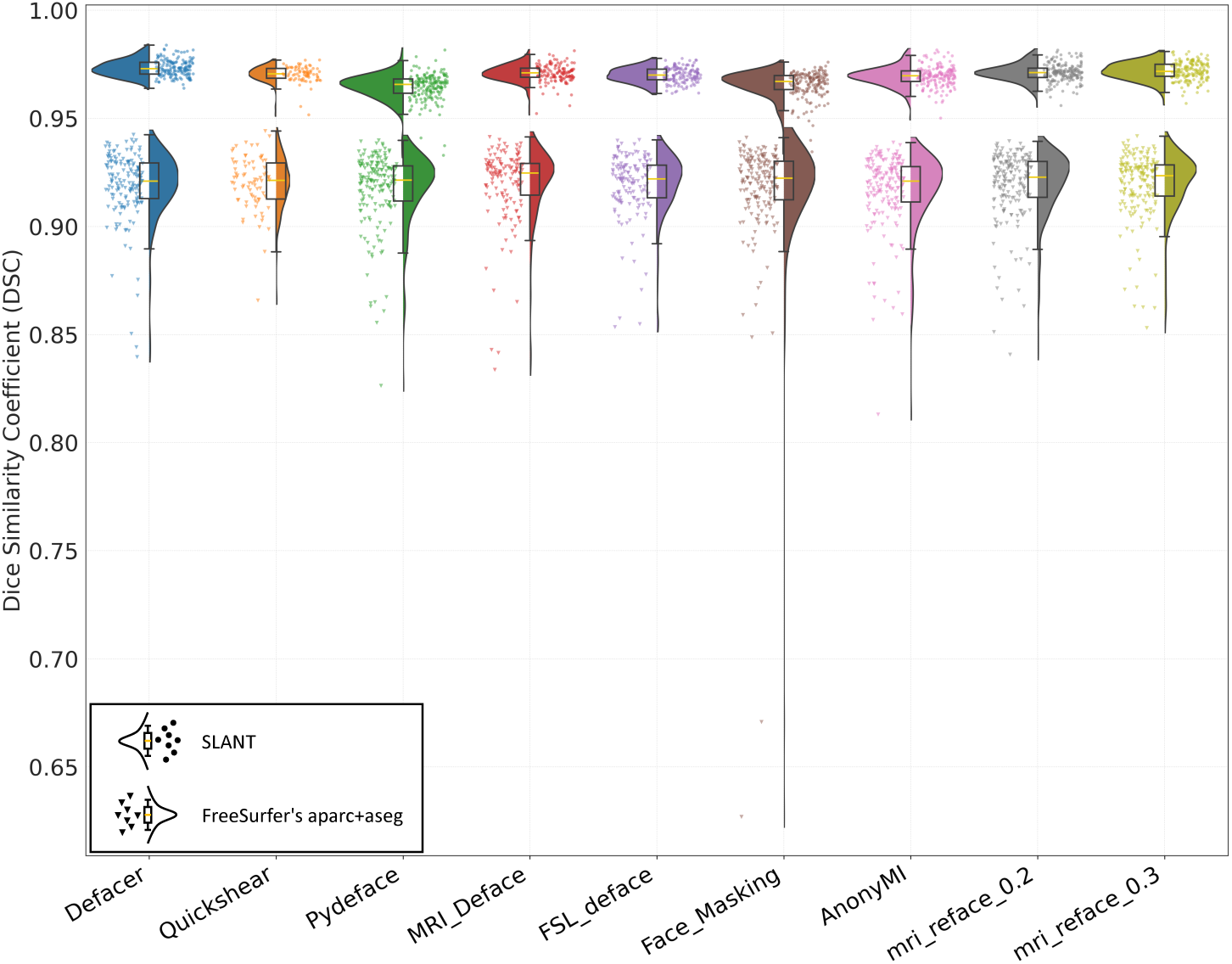
Dice Similarity Coefficient (DSC) between the segmentations of the unaltered images and the defaced images in the OASIS-3 cohort: In each column, we present the results for a specific defacing algorithm with two “raincloud” plots. The raincloud plots with the “cloud” on the left correspond to the SLANT comparison, while the plots with the “cloud” on the right correspond to FreeSurfer. The individual “raindrops” correspond to the mean DSC of the labels (by SLANT or FreeSurfer) of a specific subject from the OASIS-3 cohort.

Presenting the mean DSC per subject, as in Fig. 2, offers an incomplete picture of the performance of SLANT and FreeSurfer. Unfortunately, due to the large number of labels provided by both SLANT and FreeSurfer it is difficult to present all the results in this manuscript. As a compromise, we present DSC for some representative regions of interest (ROIs) for both SLANT and FreeSurfer in Fig. 3 and include results for all available labels in the Supplemental Material. We note that the ROIs defined by SLANT and FreeSurfer differ slightly, as they were developed from different atlases that contain different ROIs. Nonetheless, for Fig. 3 we have chosen anatomically comparable ROIs that share enough similarities for an informed side-by-side comparison.

**Fig 3:**
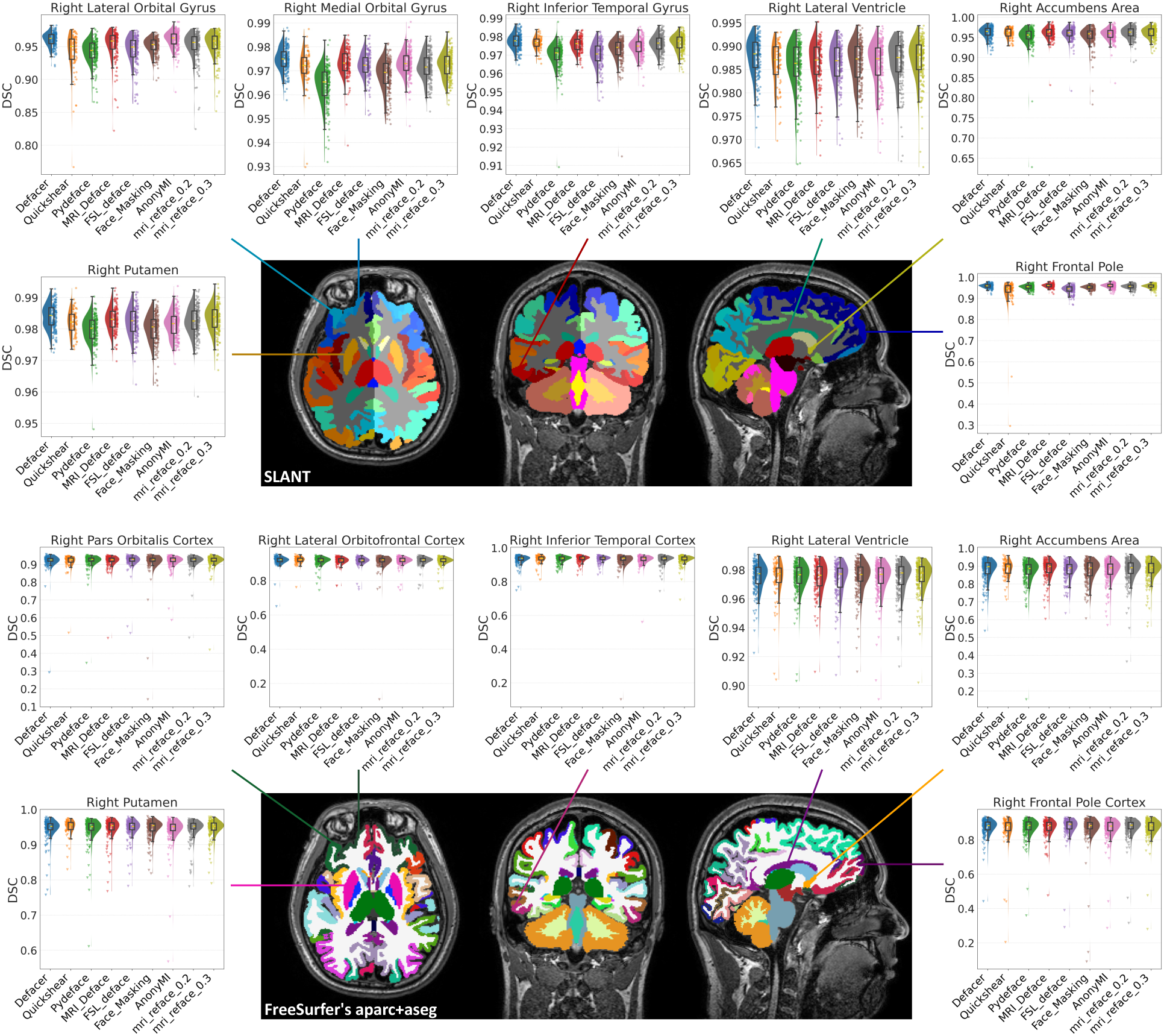
Dice Similarity Coefficient (DSC) for the segmentation (of SLANT or FreeSurfer) between unaltered and defaced images for seven regions of interest (ROIs) for subjects from the OASIS-3 cohort: The top collection of images shows SLANT labels on a particular subject from the OASIS-3 cohort. Surrounding the MRI are seven raincloud plots that correspond to specific ROIs. The bottom collection of images shows the FreeSurfer labels for the same OASIS-3 subject and raincloud plots for anatomically comparable ROIs.

The *y*-axis in each subplot covers the range of the DSC for that particular label. Refraining from setting uniform *y*-axis limits for all subplots may hinder the comparison between ROIs, but it does allow for a clearer view of the distribution of the DSC for each of the included labels. We observe that FreeSurfer results have a large range and more outliers than those of SLANT. The more extreme FreeSurfer outliers indicate instances of dramatic region specific disagreement. In Sec. 5, we provide examples of such outliers and discuss these results.

### 4.3 Kirby-21

Our second experiment is focused on the Kirby-21^39^ dataset. We first compare the performance of SLANT and FreeSurfer on the unaltered images with their performance on the defaced images. This portion of the experiment is similar to our first experiment on the OASIS-3 cohort (see Sec. 4.2) except with a smaller population size. This allows us to demonstrate the consistency of the previously observed behaviors across two different datasets. Given the rescan images in the Kirby-21 dataset, we can ascertain if the effects of defacing are comparable to that of scan-rescan differences. This is the second comparison we explore in this experiment. Both of these comparisons are presented in Fig. 4, using a visualization identical to that of Fig. 2. To prevent confusion, we separate the results from the defacing algorithms and the results from the “rescan and registration” component with a vertical line.

**Fig 4:**
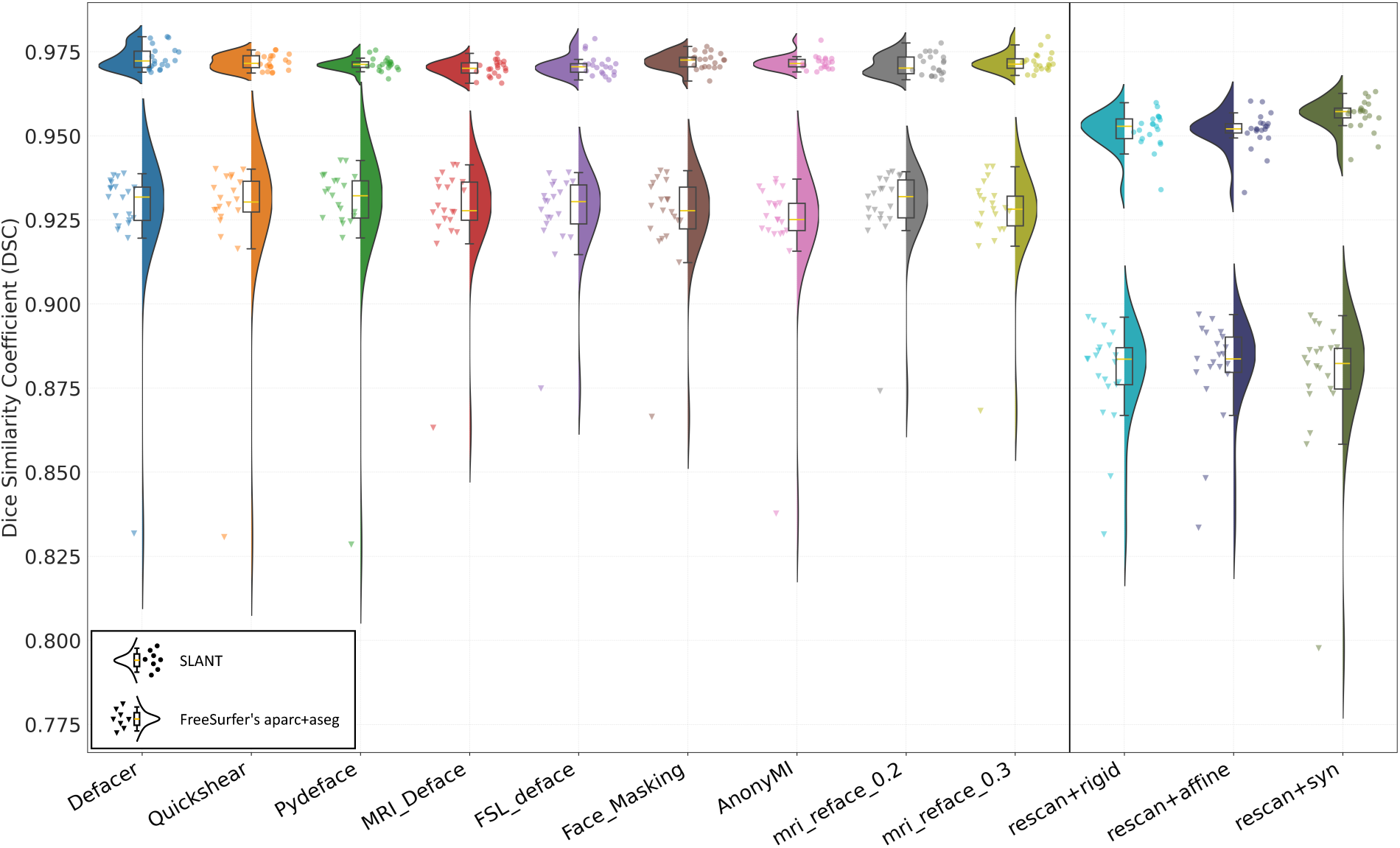
Dice Similarity Coefficient (DSC) between segmentations of the unaltered first scan, the defaced first scan, and aligned rescan on the Kirby-21 dataset: In each column, we present the results for the segmentation comparison between the unaltered first scan and either a defaced first scan or the unaltered rescan aligned to the first scan. The raindcloud plots are explained in Fig. 2. Key: “rescan+rigid” – rescan registered with rigid registration; “rescan+affine” – rescan registered with affine registration; “rescan+syn” – rescan registered with SyN based deformable registration.

Despite the smaller sample size of the Kirby-21 (*N* = 21) it exhibits a very similar trend for the defacing algorithms—left side of Fig. 4—as that of the OASIS-3 (*N* = 179) cohort as shown in Fig. 2. For the “rescan and registration” component, shown on the right side of Fig. 4, we observe that the groups are significantly lower than those of the defacing algorithms. Specifically, for the SLANT segmentation method, the DSC for the “rescan and registration” results are approximately 0.018 lower than those for the defacing methods. While for the FreeSurfer segmentations, the difference is even greater, with the DSC for the “rescan + registration” group being about 0.039 lower than the defacing methods. These findings suggest that rescanning followed by registration may have a greater impact on brain segmentation consistency than defacing alone. Moreover, these results suggest that FreeSurfer may be more susceptible than SLANT to variation from “rescan + registration”. More detailed discussion of these results is provided in Sec. 5.

## 5 Discussion

### 5.1 Quality Check Outcomes

Figure 5 presents examples of Type II Failure cases described in Sec. 3.1, where a certain proportion of the brain is removed due to excessive defacing. In the figure, we see the columns from left to right show the unaltered data and the results of *FSL_deface*, *MRI_Deface*, and *Quickshear*; while the rows from top to bottom are a 3D reconstruction of the MR data, sagittal, coronal, and axial views of the MR data overlaid with SLANT labels. As this subject failed the quality check, the SLANT and FreeSurfer results were not included in subsequent analyses. The failure of *Quick-shear* is readily observable as a sizeable portion of the frontal lobe has been removed through the defacing process. The errors of both *FSL_deface* and *MRI_Deface* might, as first viewing, not be as obvious; however, review of their coronal slices through the frontal lobe, and comparison with the unaltered image, readily demonstrate the removal of a portion of the brain. As important as defacing is for providing greater access to MRI databases, it is far more fundamental that those defacing algorithms do not remove any portion of the brain. As such those algorithms that had any Type II Failure—*Quickshear*, *MRI_Deface*, *FSL_deface*, and *Face_Masking*—are particularly worrisome. It might be argued that the Type II Failure cases of *Quickshear* can be addressed with better quality skull-stripping; this is not satisfactory, because if we have to do some high quality skull-stripping and manual review to ensure accurate brain masks, then why not release the skull-stripped images instead of the Quickshear-defaced images. This requirement of *Quickshear* is a hindrance to its adoption in many settings. The failures of *MRI_Deface* (20.1% of OASIS-3), *FSL_deface* (20.7% of OASIS-3), and *Face_Masking* (1.7% of OASIS-3) are disappointing and highlight that any use of defacing algorithms should be followed by a quality check to establish no such errors. We note that *FSL_deface* was used in the original processing of the UK BioBank^21^ data and, unfortunately, the UK BioBank does not report any manual review of these defaced images. Finally, we recall that Type I Failure cases are when the defacing algorithm failed to deface the underlying image. Such cases occurred with *Defacer* (8.9% of OASIS-3) and *MRI_Deface* (3.9% of OASIS-3), thus further highlighting the necessity of manual review of the defaced output.

**Fig 5:**
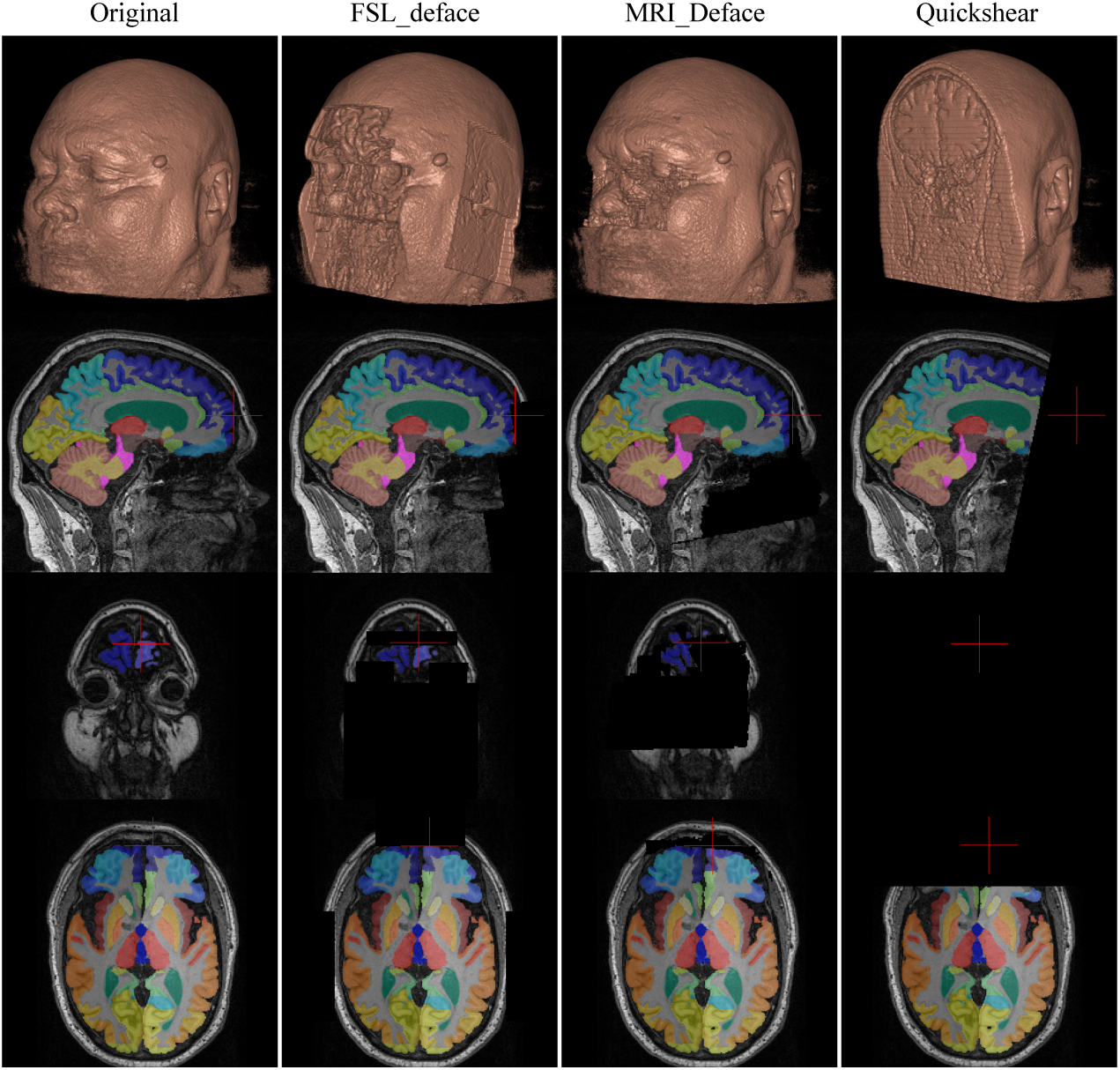
Example of Type II Failure cases: The columns from left to right show the unaltered (Original) data and the results of *FSL_deface*, *MRI_Deface*, and *Quickshear*. The rows from top to bottom show 3D renderings of the head before and after defacing by the three algorithms, then sagittal, coronal, and axial slices with their corresponding SLANT segmentation overlaid. The red cross marks the same position in each image and shows where brain voxels are removed by defacing.

### 5.2 Success and failure cases of defacing

As we note in Sec. 5.1, reliable defacing algorithms need to remove facial features effectively and ensure that the entire brain is left intact without causing any damage. Although we did not specifically evaluate the protection provided by defacing algorithms against facial recognition, we analyzed the success and failure rates of each algorithm. Of the defacing algorithms examined in this study, five of them exhibited instances of failure. Of these five, four had more than 15 instances of failure across the 200 testing samples (179 from OASIS-3 cohort and 21 from the Kirby-21 dataset). Use of these algorithms in a practical setting, requires some level of supervision, to prevent catastrophic failures such as the removal of half of the frontal lobe due to excessive defacing—see Fig. 5 for an example. However, this poses a significant challenge for researchers who need to apply defacing to large datasets since it can be time-consuming to ensure quality checks. *AnonyMI*, *Pydeface*, and both versions of *mri_reface* did not encounter any failure cases during our experiments. Nevertheless, it is important to acknowledge that passing our quality check does not guarantee successful defacing, as our definition of “success” only verifies that the algorithm processes the MR image as intended, despite the possibility of some facial voxels that were meant to be removed remaining in the image. For example, from Fig. 1 it is clear that some facial features, such as eyes and ears, were not completely removed after using *Pydeface* for defacing.

### 5.3 FreeSurfer Outliers

In this section, the FreeSurfer outliers we observe in Fig. 2 for the OASIS-3 cohort and Fig. 4 for the Kirby-21 dataset are further explored. These outliers represent the most disagreement between FreeSurfer run on the unaltered data versus being run on a defaced image. We show a specific example of an outlier from the Kirby-21 dataset in Fig. 6. In the image we show axial, coronal, and sagittal views of the FreeSurfer segmentations overlaid on the MR image for the original (unaltered) data, the data processed by the defacing algorithms, and also the registered rescan results which we only have for the Kirby-21 dataset. Each subimage features an arrow highlighting the same location in each view; we focus on this particular point as the labels assigned by FreeSurfer on the unaltered data differ dramatically with the processed data. In using the term “processed”, we are referring to the application of either a defacing algorithm or the registration of the rescan image followed by FreeSurfer. In the original image, the location is labeled as the right post-central gyrus (“ctx-rh-postcentral”). However, in the processed data, the segmentation label for this same location is consistently right pre-central gyrus (“ctx-rh-precentral”). It is worth noting that this difference in segmentation labels occurred regardless of which processing method was applied, indicating that the label for this region is highly sensitive to any processing. In Fig. 7, we present FreeSurfer segmentations corresponding to the two “worst” outliers of *Face_Masking* shown in Fig. 2, with DSCs below 0.7. Although defacing did not damage any brain voxels, the segmentation of the defaced image presents a problem. A large proportion of one hemisphere of the brain is almost unlabeled, with some crushed-glass-like labels scattered around. The cause for such failure is unclear and we note that these are the worst examples among segmentations from images that passed our defacing quality check (see Sec. 5.1). Our manual quality check does remove images that have been damaged by the defacing processing, however it clearly does not indicate that subsequent processing will be accurate.

**Fig 6:**
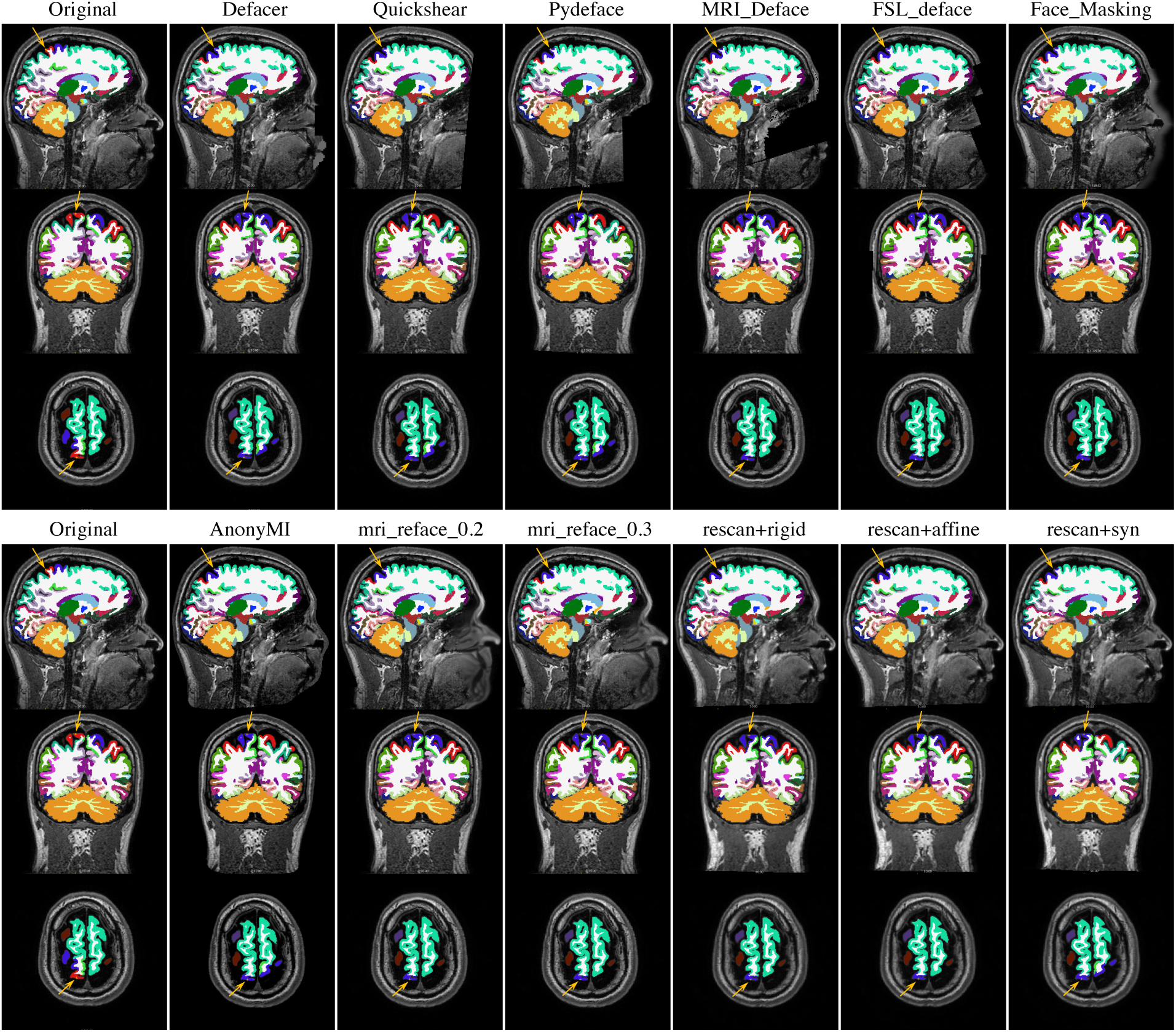
FreeSurfer Outlier Comparison: MRIs overlaid with their corresponding FreeSurfer segmentations. The arrows point to a location where the label given by FreeSurfer segmentation changed dramatically after processing either by a defacing algorithm or the registration of the rescan image. We repeat the original FreeSurfer results on both the top and bottom left column for easier comparison across the rows.

**Fig 7:**
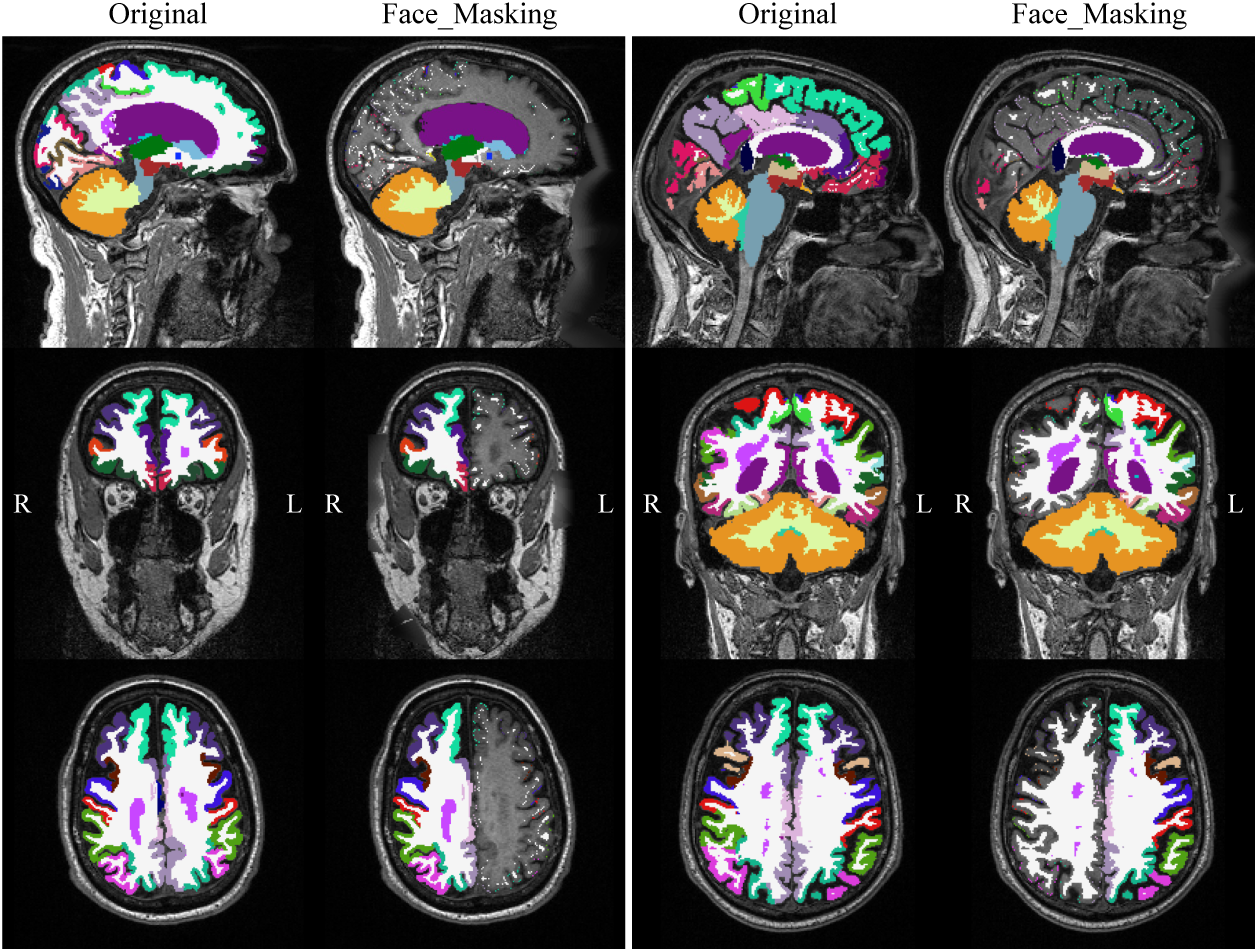
Two FreeSurfer Outliers: MRIs overlaid with their corresponding FreeSurfer segmentations. The left two columns are results from unaltered (Original) MRI and defaced by *Face_Masking* for one subject, and the right two columns are from another subject. These are the worst two subjects in our comparison with mean DSC below 0.7. Key: “L” denotes left and “R” denotes right.

### 5.4 Effects of defacing on brain segmentation

We used Dice Similarity Coefficient (DSC) to quantify the differences between the segmentation results obtained from the defaced MR images and the unaltered MR images. Figure 2 shows that both SLANT and FreeSurfer segmentations obtained from the defaced MR images differ from those obtained from the original MR images, suggesting that defacing has a potentially detrimental impact on brain segmentation. Moreover, the degree of this impact varies slightly across different defacing methods, with a stronger effect observed on FreeSurfer segmentation than on SLANT segmentation, as evidenced by the consistently lower distribution of DSC and the number of outliers for FreeSurfer. Notably, the outliers with low DSC typically represent dramatic changes in the segmentations, one of which is shown in Fig. 6, where a significant portion of the label for the right post-central gyrus (“ctx-rh-postcentral”) shifts after defacing. This shift occurs across all defacing methods and even in the rescanning group, indicating that the segmentation of this region by FreeSurfer is highly sensitive to changes in the MR image. Another, more extreme case is shown in Fig. 7, where the segmentation failed to label almost the entire hemisphere of the brain after defacing, resulting in the lowest DSC among all outliers.

For SLANT segmentation, the removal or alteration of the face affects the affine registration to the MNI atlas, the N4 bias field correction, and the intensity normalization. Additionally, as the input patches around the face are different from the original, the neural network outputs also differ. As SLANT aggregates the outputs of multiple neural networks to generate the final segmentation label image, these differences are reflected in the final results. Given that SLANT breaks down an MR image into 27 smaller regions for processing, it is then somewhat surprising that the effects of defacing can be observed in regions that are “far-away” from the changed facial features—such as changes in the occipital lobe, see Supplemental Material for an example. FreeSurfer segmentation involves registrations of multiple atlases and mapping of labels from these atlas spaces. When the facial voxels are removed or changed, the resulting registered image will differ, unless flawless and perfectly-matching brain masks were used to focus the registration on the brain, which is not feasible. Consequently, the labels mapped from the atlases can—and do—end up in different locations.

According to Fig. 4, the DSC between brain segmentations from registered rescans and the unaltered first scans are lower than the DSC between segmentations from defaced images and the original (unaltered) images. This suggests that the changes induced by rescanning a subject have a greater impact on brain segmentation than defacing alone, in general. However, this does not necessarily imply that the impact of defacing is negligible. Instead, it only provides a basis for comparison that helps us understand the magnitude of the impact. There are multiple factors that contribute to the effects of rescanning followed by alignment via registration. One factor is the light geometric deformation of the brain that occurs between the first scan and the rescan, which can happen due to the subjects being repositioned in the magnetic field. Another factor is the interpolation that occurs during the registration, which may also impact the segmentation.

### 5.5 Recommendations

We cannot overstate the importance of manual review of defacing algorithms; it is particularly critical if the data is being made available for public dissemination. Our current recommendation for a preferred defacing algorithm is *mri_reface* (version `0.3`) for the following reasons:

i. It is a user-friendly application that is readily incorporated in existing scripting pipelines;
ii. It exhibits a DSC which is slightly above average for the segmentation results between the unaltered and defaced images;
iii. It is robust to low image quality and different head positions, producing consistent results that resemble unaltered MR images.
iv. By manipulating the facial features to match a template, it clearly provides a non-identifiable image.

### 5.6 Strengths and limitations of current study

We include eight defacing algorithms that cover the majority of popular choices used in the past 15 years. To ensure that the processing steps were performed accurately, we have communicated with the authors of some of these defacing algorithms, including *mri_reface*,^25^ *AnonyMI*,^24^ and *Defacer*.^23^ We include two pipelines to analyze the effects of defacing on brain segmentation. One pipeline, SLANT, is based on deep-learning, while the other, FreeSurfer, is a widely popular multi-atlas based approach. The inclusion of these two pipelines allows for a more comprehensive comparison of the results obtained. For our analysis, we have used a total of 200 MR images and performed a quality check aimed to ensure the reliability of the results.

Due to the scope of this study—200 subjects, multiple defacing algorithms, and two neuroimaging pipelines—we only investigate the effects of defacing on brain segmentation and used only DSC to quantify these effects. Although DSC provides information on the extent of overlap between segmentation labels, it does not consider other aspects such as shape, topology, or the connected components. We did not study volumetric changes, which is a common downstream task in medical image analysis. This adds to the limitations of this study. For a more comprehensive comparison, additional metrics and analyses should be included.

As we ran the defacing algorithms on different servers with varying hardware and operating systems, we do not report the run time of each algorithm or its memory usage. As a result, we were unable to provide evidence for questions regarding algorithm efficiency. Furthermore, we did not provide evidence to support our recommendation of *mri_reface*. We also did not measure the effectiveness of the defacing algorithms in protecting against identification, which is a crucial aspect to consider when selecting a reliable defacing algorithm.

## 6 Conclusion

Defacing MR images has effects on brain segmentation. While the effects, quantified using DSC, are less than those of rescanning followed by registration, they are still noticeable and should not be disregarded. In particular, it is important to pay extra attention to the possibility of catastrophic failures of brain segmentation caused by defacing. In the worst scenario, brain voxels can be removed due to excessive defacing by some algorithms. To prevent this problem, a thorough quality check is necessary before using defaced images. Using robust algorithms, which in our experience were *mri_reface* and *AnonyMI*, can alleviate the burden of manual review. There are other scenarios where the problems are less noticeable but can also have devastating effects on neuroanalysis. For instance, the output of a segmentation pipeline for a specific brain region can be highly sensitive to changes in MR images. Or, in some extreme cases, the segmentation pipeline malfunctions on the entire hemisphere of the brain in the defaced MRI, and fails to output any labels–see Fig. 7 for example. To address these issues, it can be helpful to use multiple segmentation pipelines, especially those that are more invariant to changes of non-brain voxels in MR images, in order to draw reliable conclusions.

### Disclosures

No conflicts of interest.

### Code, Data, and Materials Availability

The datasets used in this study are open-access and can be accessed through straightforward online applications. All software packages used in this study are publicly available. We provide the URLs for the datasets and software packages in the Supplementary Material. For researchers interested in reproducing our analysis results, we also list the sample subjects from the datasets in the Supplementary Material.

## Data Availability

All data used for this study are available online at:

https://oasis-brains.org/

https://www.nitrc.org/projects/multimodal/

https://oasis-brains.org/

https://www.nitrc.org/projects/multimodal/

## Acknowledgments

This work was supported by the National Institute of Health through National Institute of Neurological Disorders and Stroke grant 1R21NS120286-01 (PI: J.L. Prince) and the National Institute of Biomedical Imaging and Bioengineering grant 1R01EB017230-01A1 (PI: B.A. Landman).

**Chenyu Gao** is a Ph.D. student in electrical and computer engineering at Vanderbilt University, working with Prof. Bennett Landman on the harmonization of diffusion MRI. His research interest is focused on image processing and computer vision with application to medical image analysis. He received his BS degree in biomedical engineering from Sun Yat-sen University, and his MS degree in biomedical engineering from Johns Hopkins University, working with Prof. Jerry Prince.

Biographies and photographs of the other authors are not available.

## Footnotes

1 139 per 1,000 in 2016 would have equated to 44 million scans.

2 https://github.com/MASILab/SLANTbrainSeg

