## Supplementary Material for "A reproducibility evaluation of the effects of MRI defacing on brain segmentation"

**Chenyu Gao**[a,*], **Bennett A. Landman**[a], **Jerry L. Prince**[b], **Aaron Carass**[b]
[a]Vanderbilt University, Department of Electrical and Computer Engineering, Nashville, Tennessee, 37235
[b]The Johns Hopkins University, Department of Electrical and Computer Engineering, Baltimore, Maryland, 21218

### S1 Defacing Algorithms

Here we outline the eight defacing algorithms included in our experiments.

#### *Defacer [1]*

To our knowledge, *Defacer* is the first open-source algorithm published in the literature that uses a deep-learning based method for MRI anonymization. Two hundred and forty whole head MRIs from the Alzheimer's Disease Neuroimaging Initiative (ADNI) [2] database were manually labeled and used to train a 3D attention-gated U-net based network. To overcome issues with the small sample size of the training data, the authors used extensive data augmentation techniques at each training epoch. The task of the network is detection of the eyes, ears, nose, and mouth in an MR image. After detection, the anonymization process proceeds by using straightforward image processing techniques to manipulate the intensity values of the detected facial feature voxels and their immediate surroundings. The authors validate their performance by using a test set of one hundred whole head MRIs from the Open Access Series of Imaging Studies (OASIS) [3] database. We use the version cloned from the authors' GitHub repository that was updated on November 19th, 2020.

#### *Quickshear [4]*

*Quickshear* defaces by identifying a plane that divides the whole head MRI into two sections, one containing facial features and another containing the entire brain. Those voxels that are deemed to be on the side of the plane containing the face are set to be zero; which the authors describe as "effectively shearing off identifiable facial features." To achieve this *Quickshear* requires a brain mask as input, from which it creates an edge of the brain collapsed to the sagittal plane. From this 2D projection an edge-of-the-brain mask is generated and the convex hull of which provides the line of separation between facial and brain voxels in the mid-sagittal plane. This 2D line of separation is replicated in each sagittal slice creating the "shearing plane" which denotes the boundary between the front of the head and the brain. The space between the "shearing plane" and the provided brain mask is a tunable parameter. *Quickshear* was originally evaluated on scan-rescan data from the Kirby-21 dataset [5], with the quantification being the consistency of the defacing. We use version `1.1.0` released on March 23rd, 2017. The brain masks required by *Quickshear* were generated by BET (Brain Extraction Tool) [6] with default settings.

*MRI_Deface [7]*

*MRI_Deface* performs a linear registration [8] between a template image and an input image. The template has a corresponding atlas of face memberships that was manually created by labeling the facial features of ten subjects. The facial features comprise the entire front of the head. A brain mask is constructed of those voxels that have a non-zero probability—based on the template registration—of being within the brain. This binarized mask undergoes several morphological operations, to ensure no brain voxels are left behind. All voxels that are outside the mask and have a non-zero probability of being a facial feature have their intensity set to 0. The authors specify that their approach should only be used with $T_1$-weighted MRIs. The authors include an evaluation on the downstream task of skull-stripping, to demonstrate the stability of their method. We use version `1.22`, with default settings.

*Pydeface [9]*

*Pydeface* is a popular tool for MRI defacing among the Python neuroimaging community, due to its relative ease of installation and use. It is similar to *MRI_Deface*, in that it comes with its own template atlas which is registered using FLIRT [10] to an input image. *Pydeface* then applies the template masks to the input image to remove facial voxels. *Pydeface*, unlike *MRI_Deface*, does not perform any morphological operations to the transformed facial mask. We use version `2.0.0`, with default settings.

*FSL_deface [11]*

*FSL_deface* follows the same workflow as *MRI_Deface* and *Pydeface*, in that it registers its template and corresponding masks to the input image. Like *Pydeface* it uses FLIRT for the linear registration step and does not perform any morphological operations on the transformed mask. The authors take the additional step to compute the overlap between non-defaced brain masks and the defacing masks with only 0.5% of their reported results containing any overlap. The authors performed a manual review of their overlap cases and all were deemed acceptable. *FSL_deface* has one other key difference with *MRI_Deface* or *Pydeface*, it includes the ears in its defacing mask. Given the identifiability of the ears [12], this seems like an unfortunate oversight of *MRI_Deface* and *Pydeface*. The authors report that *FSL_deface* can be used on both $T_1$- and $T_2$-weighted MRIs. We use the version released with FSL `6.0.3`, with default settings.

*Face_Masking [13]*

*Face_Masking* (or "Normalized Anterior Filtering" as it was called in its original paper) rather than simply masking the face, as its name implies, focuses on blurring the facial surface instead. In doing so it is designed to avoid impinging on areas of interest and also does not introduce hard intensity edges to the whole head MRI that can confuse subsequent processing tools. *Face_Masking* extracts a thin boundary layer containing facial anatomy from a region of interest. This boundary layer is then stretched and flattened to fit into a box-like volume. The box-like volume is then smoothed along a plane parallel with the facial surface and then transformed back to the original data. The result is an artificial *cubist*-like face. The authors demonstrate their approach on MR and computed tomography scans and include a study of the effects of *Face_Masking* on skull-stripping and bias-field correction. We use version `10.15.2018` in a Matlab Runtime Environment (MathWorks, Natick, MA.) on a Linux-x86_64 machine.

*AnonyMI [14]*

*AnonyMI* consists of three main steps. First a water-shed algorithm [15] is applied to obtain a 3D reconstruction of the outer surface of the head (ie. skin) and the skull. Second the location of the face and ears are identified using non-linear registration between the input image and the IXI ANTs template [16, 17]. The landmarks contained in the atlas image are transformed to the input image. Finally, a defacing mask is created for the input by taking the intersection of the landmarks with the 3D skin reconstruction from the first step. These intersection areas are filled with random numbers, having the same intensity distribution as the voxels between the 3D reconstructions of the skin and skull; values outside the skin surface are set to zero. We use the version cloned from the authors' GitHub repository that was last updated on November 11$^{th}$, 2022, as a plug-in to 3D Slicer (version `5.0.3`), with default settings.

*mri_reface [18]*

Rather than removing facial voxels, *mri_reface* replaces those voxels with a population average face created from 3D $T_1$-weighted MPRAGE images from 177 Mayo Clinic participants aged 30–89. *mri_reface* does this by first registering their template with the input image using ANTs [16]. They then apply the deformation field to the average face template, which preserves the original brain data but replaces the facial features with those of the template. To help blend the new facial features, *mri_reface* transforms the image intensities of the template image to match those of the input image. This is done using a combination of global and piece-wise intensity matching—similar to [19]—followed by bias correction for smooth local intensity normalization between the images [20]. In our previous study [21], we utilized version `0.2` of *mri_reface*. In the present study, we incorporate both version `0.2` and the recently released version `0.3` of *mri_reface*. Version `0.3` has new features that make the resulting re-faced images appear more natural. We note that [18] evaluated whether MR images were correctly recognizable, by facial recognition software, after defacing. Of the presented methods in that paper, the ranking in terms of correct matching between photos and MRIs of participants after defacing was: *FSL_Deface* (28%); *mri_reface* (30%); *MRI_Deface* (33%); *PyDeface* (38%). The authors also included an intra-class correlation coefficient (ICC) comparison between the presented methods before and after defacing. ICC would identify brain structures that have changed their volume in some way, it would not highlight changes in the spatial positioning of those brain structures which is critically important when potentially considering the accidental removal of portions of the brain due to defacing. We note this, as we have observed brain structures "*moving*" if their segmentation is performed on the original or defaced images, see the manuscript for an example.

### S2 Links to defacing software

`Defacer` [1] https://github.com/yeonuk-Jeong/Defacer
`Quickshear` [4] https://github.com/nipy/quickshear
`MRI_Deface` [7] https://surfer.nmr.mgh.harvard.edu/fswiki/AutomatedDefacingTools
`Pydeface` [9] https://github.com/poldracklab/pydeface
`FSL_deface` [11] https://fsl.fmrib.ox.ac.uk/fsl/fslwiki/FSL
`Face_Masking` [13] https://wiki.xnat.org/xnat-tools/face-masking
`AnonyMI` [14] https://github.com/iTCf/anonymi
`mri_reface` [18] https://www.nitrc.org/projects/mri_reface

### S3 Links to datasets

`OASIS-3` [22] https://oasis-brains.org/
`Kirby-21` [5] https://www.nitrc.org/projects/multimodal/

### S4 DSC of all brain regions

In Figs. S1 thru S3, we present DSCs of all ROIs defined by FreeSurfer segmentation. In Figs. S4 thru S6, we present DSCs of all ROIs defined by SLANT.

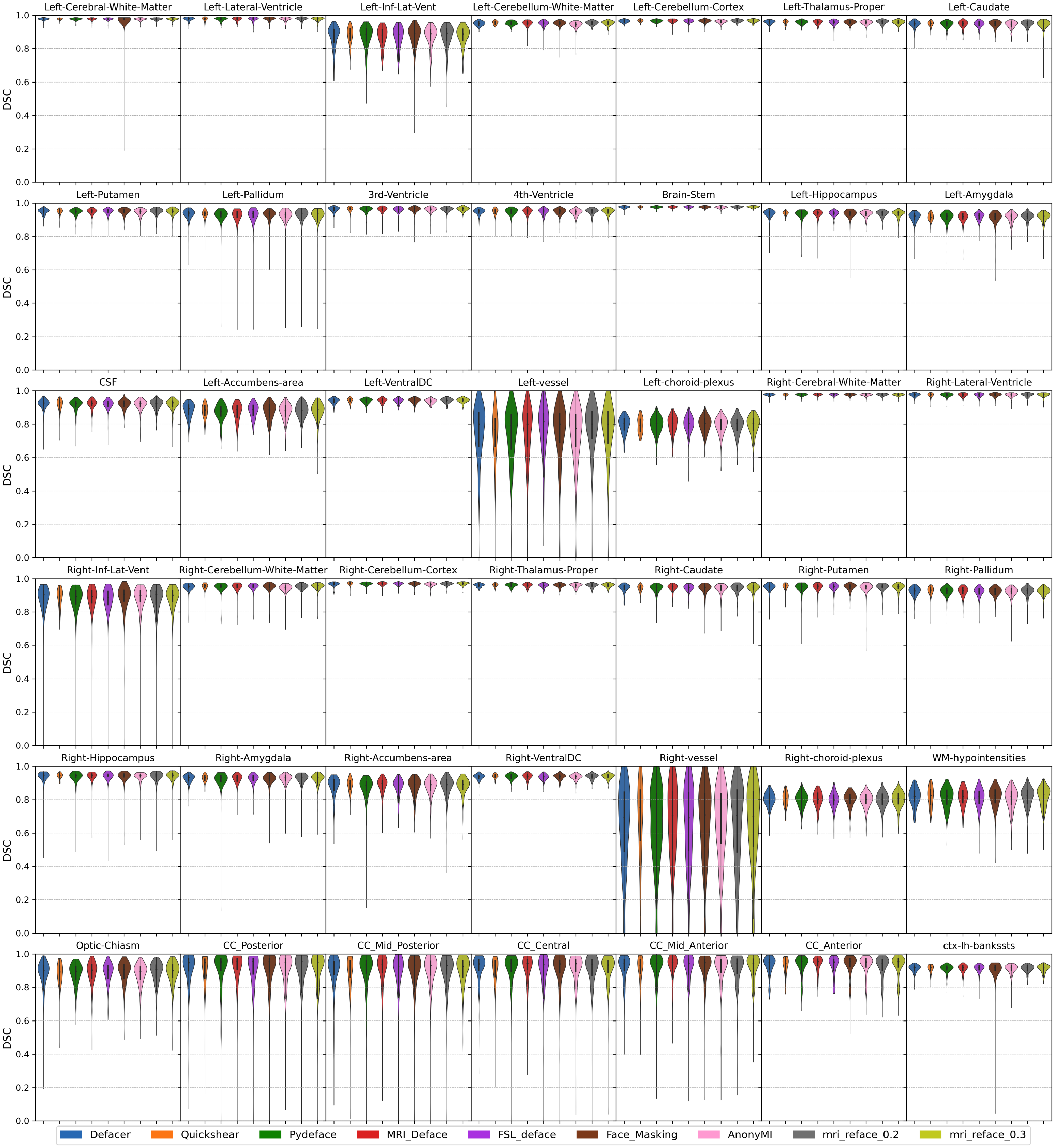

**Fig S1 Dice Similarity Coefficient (DSC) between the FreeSurfer segmentations of the unaltered images and the defaced images in the OASIS-3 cohort.** Figures S1 thru S3 include all ROIs defined by FreeSurfer.

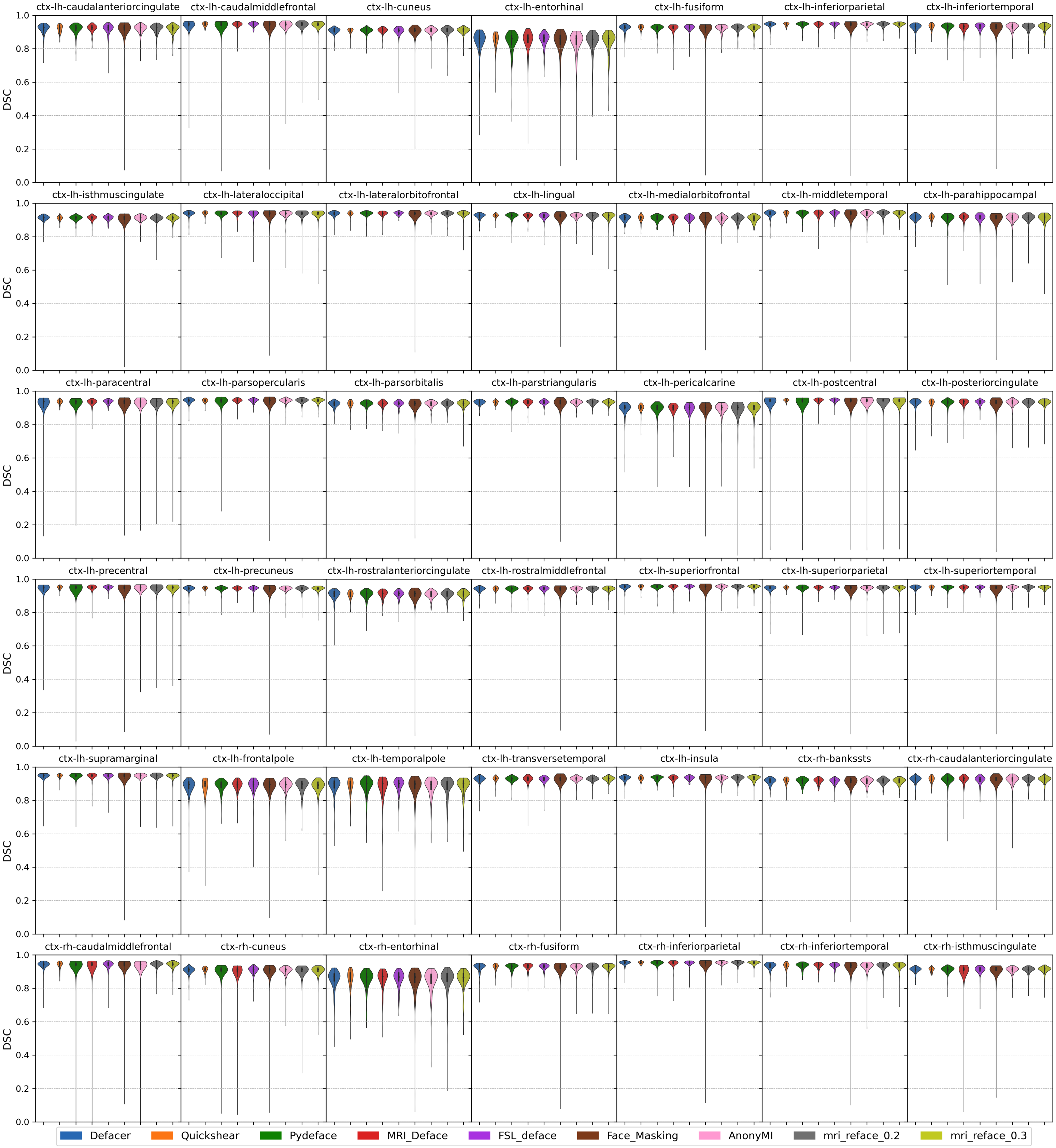

**Fig S2 Dice Similarity Coefficient (DSC) between the FreeSurfer segmentations of the unaltered images and the defaced images in the OASIS-3 cohort.** Figures S1 thru S3 include all ROIs defined by FreeSurfer.

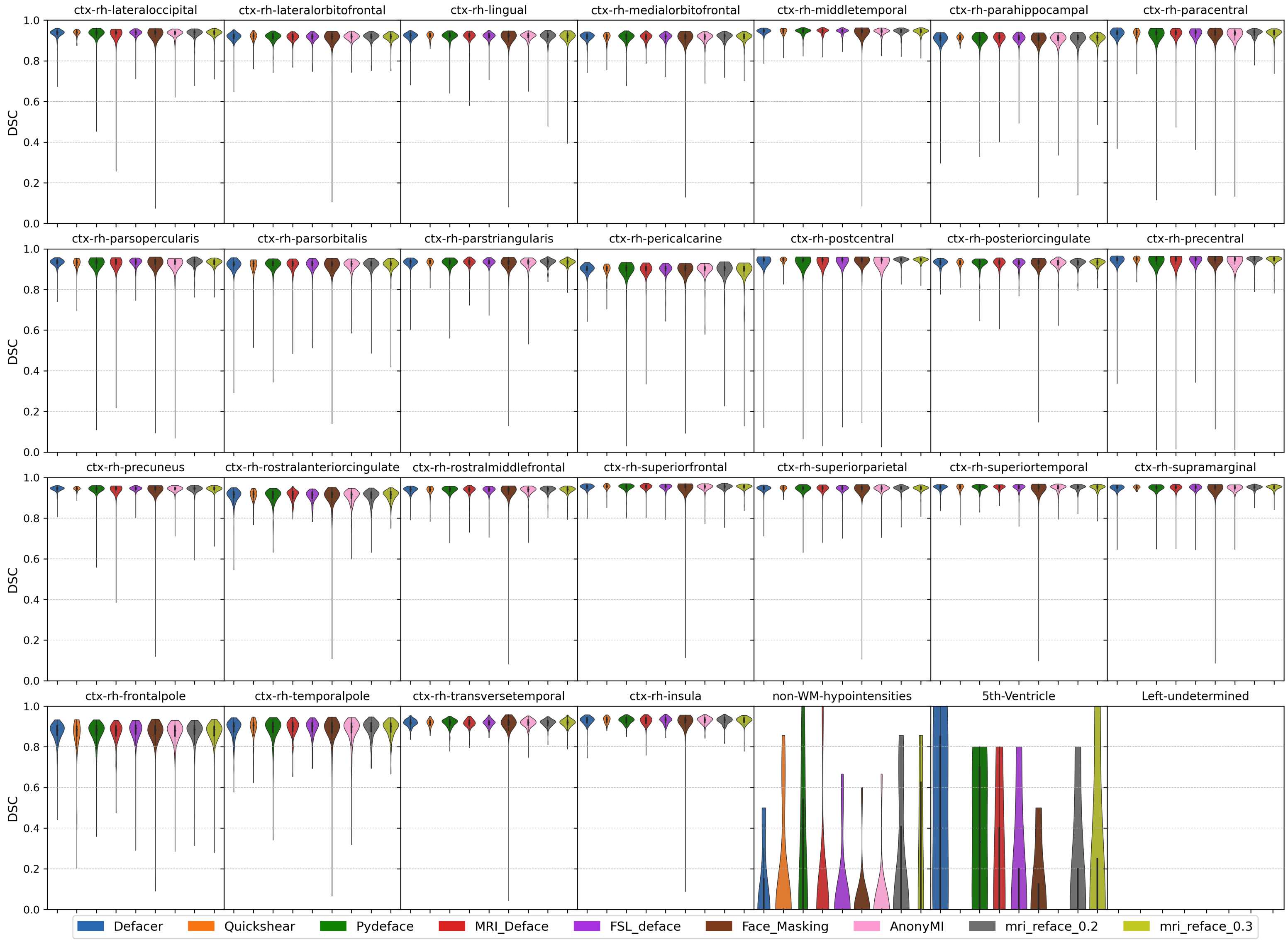

**Fig S3 Dice Similarity Coefficient (DSC) between the FreeSurfer segmentations of the unaltered images and the defaced images in the OASIS-3 cohort.** Figures S1 thru S3 include all ROIs defined by FreeSurfer.

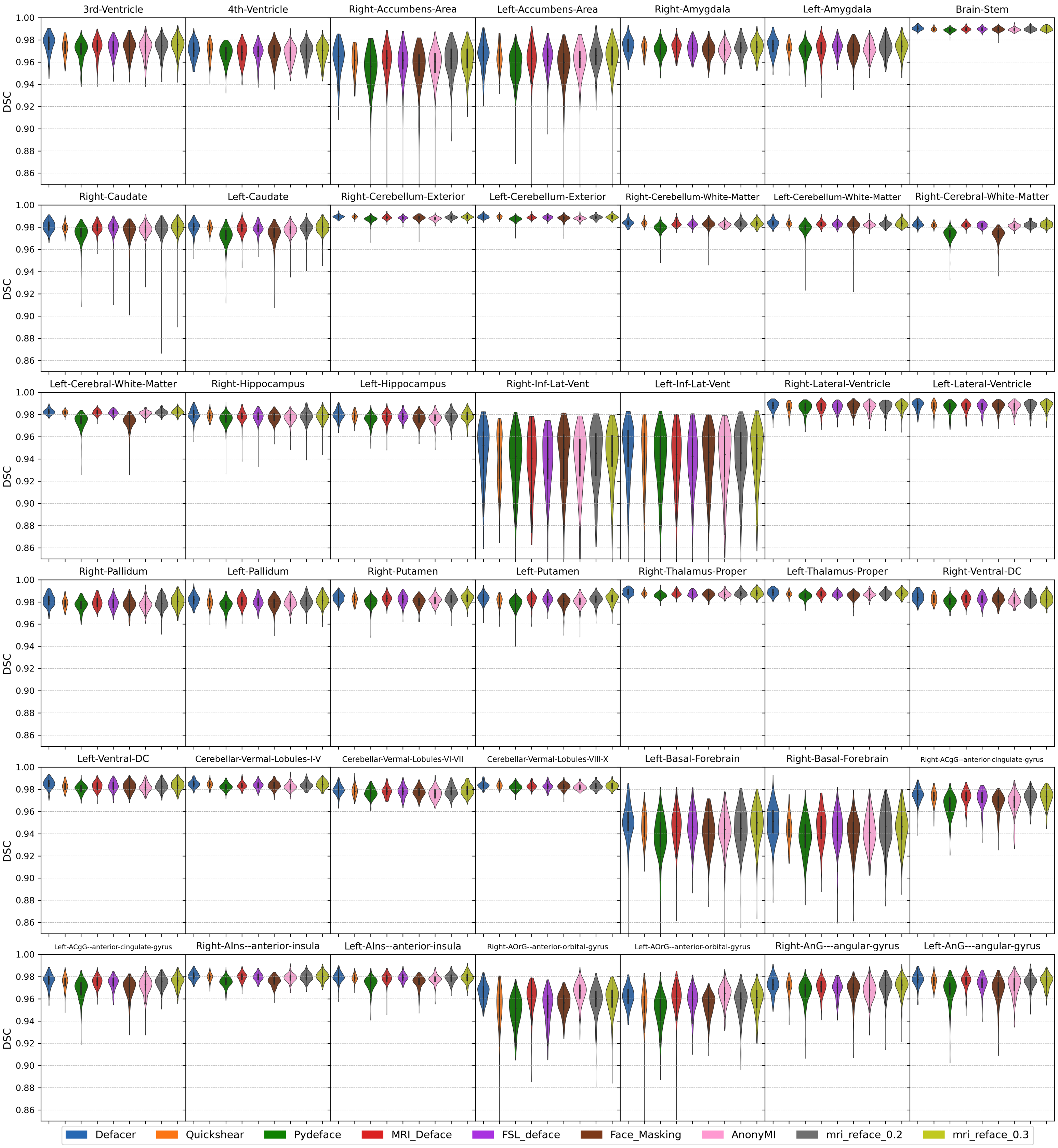

**Fig S4 Dice Similarity Coefficient (DSC) between the SLANT segmentations of the unaltered images and the defaced images in the OASIS-3 cohort.** Figures S4 thru S6 include all ROIs defined by SLANT.

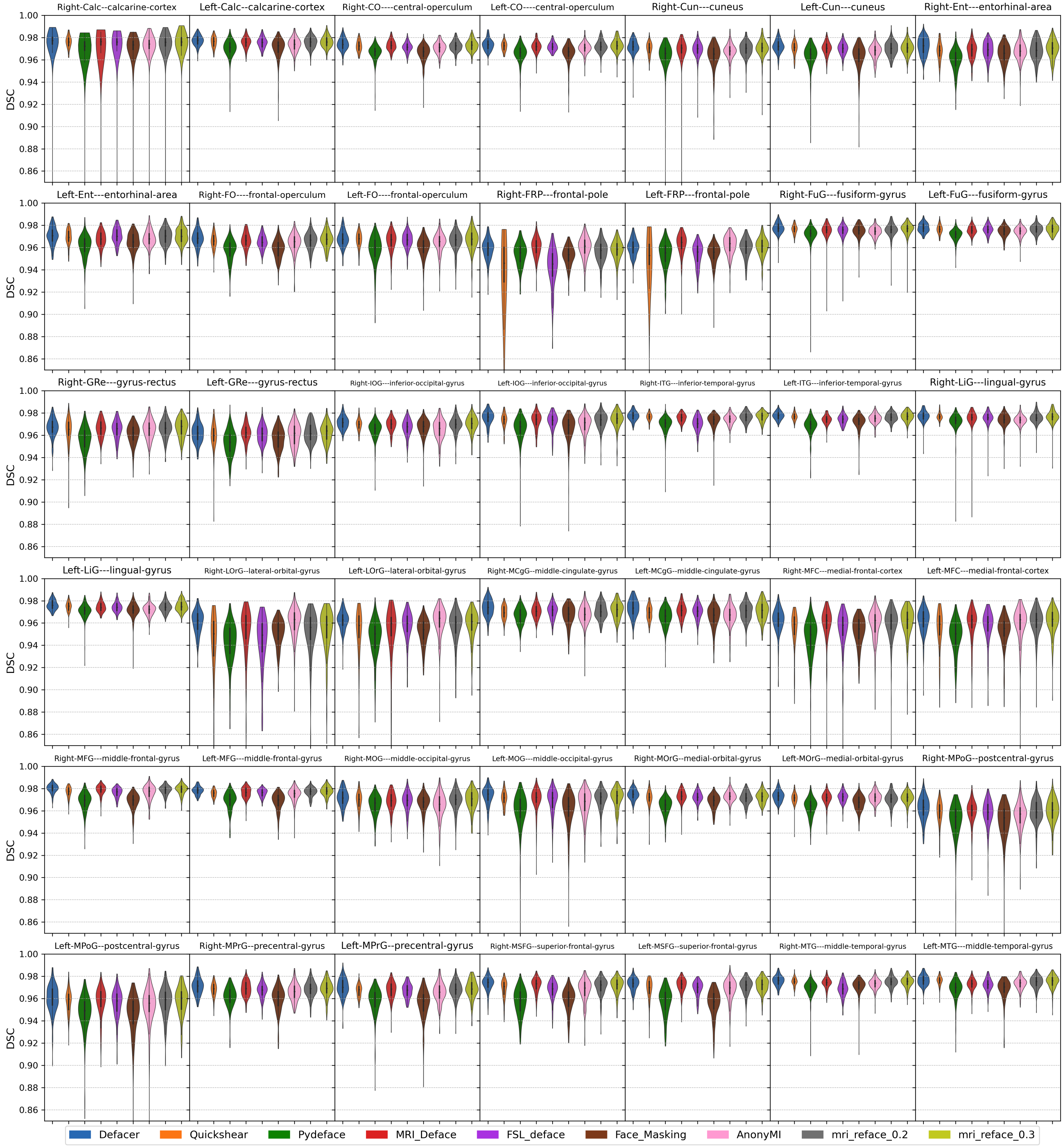

**Fig S5 Dice Similarity Coefficient (DSC) between the SLANT segmentations of the unaltered images and the defaced images in the OASIS-3 cohort.** Figures S4 thru S6 include all ROIs defined by SLANT.

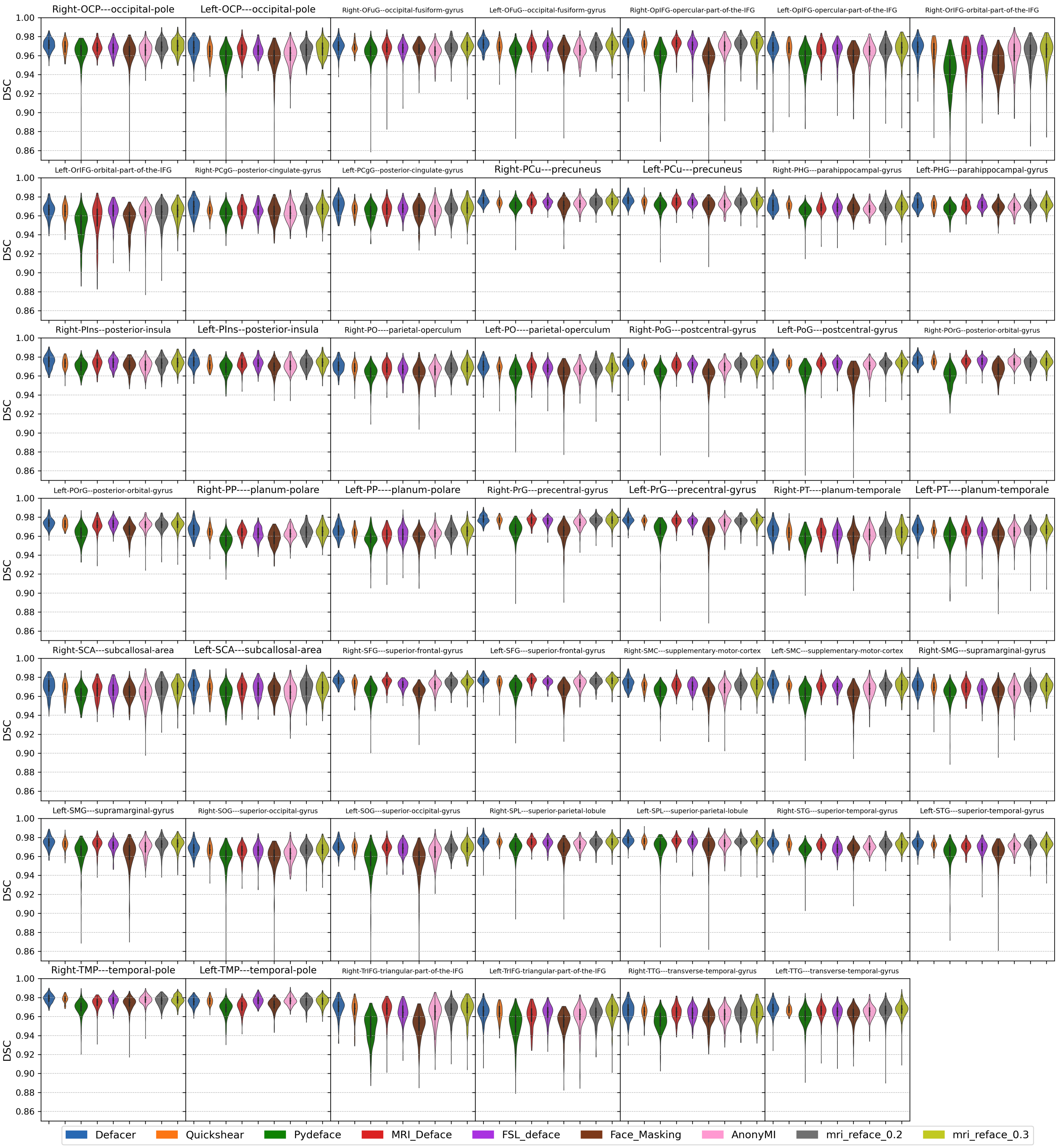

**Fig S6 Dice Similarity Coefficient (DSC) between the SLANT segmentations of the unaltered images and the defaced images in the OASIS-3 cohort.** Figures S4 thru S6 include all ROIs defined by SLANT.

### S5 FreeSurfer Outlier

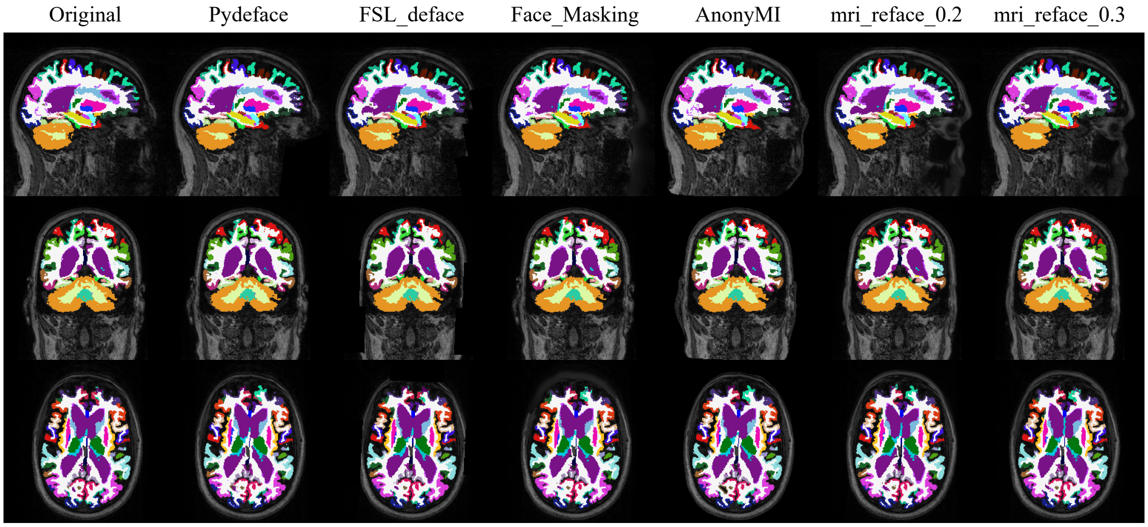

**Fig S7 Example of FreeSurfer Outlier in the OASIS-3 Experiment:** MRIs overlaid with their corresponding FreeSurfer segmentations.

### S6 Manual QA Review

In Tables S1 and S2, we present the results of the manual review of our OASIS-3 cohort and Kirby-21 dataset, respectively. See the manuscript for an explanation of Type I and Type II Failures.

Table S1: Results of the manual review of the 179 $T_1$-weighted images from the OASIS-3 cohort. **Key:** 0 – Success; I – Type I Failure;
II – Type II Failure.

| **Image** | Defacer | Quickshear | Pydeface | MRI_Deface | FSL_deface | Face_Masking | AnonyMI | mri_reface_0.2 | mri_reface_0.3 |
|---|---|---|---|---|---|---|---|---|---|
| sub-OAS30005_ses-d0143_T1w.nii.gz | 0 | 0 | 0 | 0 | 0 | 0 | 0 | 0 | 0 |
| sub-OAS30007_ses-d0061_run-01_T1w.nii.gz | 0 | II | 0 | 0 | 0 | 0 | 0 | 0 | 0 |
| sub-OAS30008_ses-d1327_T1w.nii.gz | 0 | II | 0 | 0 | 0 | 0 | 0 | 0 | 0 |
| sub-OAS30010_ses-d0068_T1w.nii.gz | 0 | 0 | 0 | I | 0 | 0 | 0 | 0 | 0 |
| sub-OAS30015_ses-d2004_T1w.nii.gz | 0 | II | 0 | 0 | 0 | 0 | 0 | 0 | 0 |
| sub-OAS30024_ses-d0084_T1w.nii.gz | 0 | 0 | 0 | II | 0 | 0 | 0 | 0 | 0 |
| sub-OAS30026_ses-d0696_T1w.nii.gz | I | II | 0 | II | 0 | 0 | 0 | 0 | 0 |
| sub-OAS30034_ses-d0044_T1w.nii.gz | 0 | 0 | 0 | 0 | 0 | 0 | 0 | 0 | 0 |
| sub-OAS30043_ses-d0145_T1w.nii.gz | 0 | II | 0 | 0 | 0 | 0 | 0 | 0 | 0 |
| sub-OAS30050_ses-d0110_T1w.nii.gz | 0 | 0 | 0 | 0 | 0 | 0 | 0 | 0 | 0 |
| sub-OAS30056_ses-d3491_T1w.nii.gz | 0 | 0 | 0 | 0 | 0 | 0 | 0 | 0 | 0 |
| sub-OAS30063_ses-d0160_run-01_T1w.nii.gz | 0 | 0 | 0 | 0 | 0 | 0 | 0 | 0 | 0 |
| sub-OAS30064_ses-d0687_run-01_T1w.nii.gz | 0 | II | 0 | 0 | 0 | 0 | 0 | 0 | 0 |
| sub-OAS30065_ses-d0548_T1w.nii.gz | 0 | II | 0 | 0 | 0 | 0 | 0 | 0 | 0 |
| sub-OAS30077_ses-d0944_T1w.nii.gz | I | 0 | 0 | 0 | 0 | 0 | 0 | 0 | 0 |
| sub-OAS30085_ses-d1566_T1w.nii.gz | 0 | II | 0 | II | II | 0 | 0 | 0 | 0 |
| sub-OAS30090_ses-d0118_T1w.nii.gz | 0 | II | 0 | 0 | II | 0 | 0 | 0 | 0 |
| sub-OAS30092_ses-d0636_T1w.nii.gz | 0 | 0 | 0 | 0 | 0 | 0 | 0 | 0 | 0 |
| sub-OAS30098_ses-d0036_T1w.nii.gz | 0 | 0 | 0 | 0 | 0 | 0 | 0 | 0 | 0 |
| sub-OAS30104_ses-d0328_run-01_T1w.nii.gz | 0 | 0 | 0 | 0 | 0 | 0 | 0 | 0 | 0 |
| sub-OAS30128_ses-d0044_T1w.nii.gz | 0 | II | 0 | II | 0 | 0 | 0 | 0 | 0 |

Table S1: Results of the manual review of the 179 $T_1$-weighted images from the OASIS-3 cohort. **Key:** 0 – Success; I – Type I Failure;
II – Type II Failure.

| **Image** | Defacer | Quickshear | Pydeface | MRI_Deface | FSL_deface | Face_Masking | AnonyMI | mri_reface_0.2 | mri_reface_0.3 |
|---|---|---|---|---|---|---|---|---|---|
| sub-OAS30140_ses-d0172_run-01_T1w.nii.gz | 0 | II | 0 | 0 | 0 | 0 | 0 | 0 | 0 |
| sub-OAS30142_ses-d0075_run-01_T1w.nii.gz | 0 | 0 | 0 | 0 | 0 | 0 | 0 | 0 | 0 |
| sub-OAS30143_ses-d3856_T1w.nii.gz | 0 | II | 0 | II | 0 | 0 | 0 | 0 | 0 |
| sub-OAS30146_ses-d3322_T1w.nii.gz | 0 | II | 0 | 0 | 0 | 0 | 0 | 0 | 0 |
| sub-OAS30150_ses-d0100_run-01_T1w.nii.gz | 0 | 0 | 0 | 0 | 0 | 0 | 0 | 0 | 0 |
| sub-OAS30151_ses-d0064_T1w.nii.gz | 0 | 0 | 0 | 0 | 0 | 0 | 0 | 0 | 0 |
| sub-OAS30163_ses-d0091_run-01_T1w.nii.gz | 0 | II | 0 | II | II | 0 | 0 | 0 | 0 |
| sub-OAS30170_ses-d0005_T1w.nii.gz | 0 | II | 0 | II | 0 | 0 | 0 | 0 | 0 |
| sub-OAS30183_ses-d0082_run-01_T1w.nii.gz | 0 | 0 | 0 | 0 | 0 | 0 | 0 | 0 | 0 |
| sub-OAS30191_ses-d2352_T1w.nii.gz | I | II | 0 | II | 0 | 0 | 0 | 0 | 0 |
| sub-OAS30208_ses-d1703_T1w.nii.gz | 0 | 0 | 0 | II | 0 | 0 | 0 | 0 | 0 |
| sub-OAS30218_ses-d0841_T1w.nii.gz | 0 | II | 0 | 0 | 0 | 0 | 0 | 0 | 0 |
| sub-OAS30220_ses-d0104_T1w.nii.gz | 0 | II | 0 | 0 | 0 | 0 | 0 | 0 | 0 |
| sub-OAS30225_ses-d0482_T1w.nii.gz | 0 | II | 0 | 0 | 0 | 0 | 0 | 0 | 0 |
| sub-OAS30229_ses-d0101_T1w.nii.gz | 0 | II | 0 | II | II | 0 | 0 | 0 | 0 |
| sub-OAS30237_ses-d2478_run-01_T1w.nii.gz | 0 | II | 0 | 0 | 0 | 0 | 0 | 0 | 0 |
| sub-OAS30238_ses-d0037_T1w.nii.gz | 0 | II | 0 | 0 | II | 0 | 0 | 0 | 0 |
| sub-OAS30240_ses-d3843_T1w.nii.gz | 0 | II | 0 | II | 0 | 0 | 0 | 0 | 0 |
| sub-OAS30249_ses-d1238_run-01_T1w.nii.gz | 0 | II | 0 | 0 | 0 | 0 | 0 | 0 | 0 |
| sub-OAS30258_ses-d1225_T1w.nii.gz | 0 | II | 0 | II | 0 | 0 | 0 | 0 | 0 |
| sub-OAS30265_ses-d0192_run-01_T1w.nii.gz | 0 | II | 0 | 0 | 0 | 0 | 0 | 0 | 0 |

Table S1: Results of the manual review of the 179 $T_1$-weighted images from the OASIS-3 cohort. **Key:** 0 – Success; I – Type I Failure;
II – Type II Failure.

| **Image** | Defacer | Quickshear | Pydeface | MRI_Deface | FSL_deface | Face_Masking | AnonyMI | mri_reface_0.2 | mri_reface_0.3 |
|---|---|---|---|---|---|---|---|---|---|
| sub-OAS30275_ses-d1006_run-01_T1w.nii.gz | 0 | 0 | 0 | 0 | 0 | 0 | 0 | 0 | 0 |
| sub-OAS30282_ses-d0040_T1w.nii.gz | I | 0 | 0 | 0 | II | 0 | 0 | 0 | 0 |
| sub-OAS30285_ses-d0055_run-01_T1w.nii.gz | 0 | 0 | 0 | 0 | 0 | 0 | 0 | 0 | 0 |
| sub-OAS30293_ses-d1221_T1w.nii.gz | 0 | II | 0 | 0 | 0 | II | 0 | 0 | 0 |
| sub-OAS30318_ses-d2975_T1w.nii.gz | 0 | II | 0 | II | II | 0 | 0 | 0 | 0 |
| sub-OAS30342_ses-d0429_T1w.nii.gz | 0 | 0 | 0 | II | II | 0 | 0 | 0 | 0 |
| sub-OAS30345_ses-d0087_run-01_T1w.nii.gz | 0 | 0 | 0 | 0 | 0 | 0 | 0 | 0 | 0 |
| sub-OAS30349_ses-d3241_T1w.nii.gz | 0 | 0 | 0 | 0 | 0 | 0 | 0 | 0 | 0 |
| sub-OAS30356_ses-d0714_run-01_T1w.nii.gz | 0 | 0 | 0 | 0 | 0 | 0 | 0 | 0 | 0 |
| sub-OAS30363_ses-d0880_run-01_T1w.nii.gz | 0 | 0 | 0 | 0 | 0 | 0 | 0 | 0 | 0 |
| sub-OAS30370_ses-d0141_run-01_T1w.nii.gz | I | II | 0 | I | II | 0 | 0 | 0 | 0 |
| sub-OAS30373_ses-d1211_T1w.nii.gz | 0 | II | 0 | 0 | II | 0 | 0 | 0 | 0 |
| sub-OAS30376_ses-d0082_T1w.nii.gz | 0 | 0 | 0 | 0 | 0 | 0 | 0 | 0 | 0 |
| sub-OAS30384_ses-d0108_T1w.nii.gz | 0 | II | 0 | 0 | 0 | 0 | 0 | 0 | 0 |
| sub-OAS30401_ses-d0106_run-01_T1w.nii.gz | 0 | 0 | 0 | 0 | 0 | 0 | 0 | 0 | 0 |
| sub-OAS30403_ses-d2758_T1w.nii.gz | 0 | 0 | 0 | I | 0 | 0 | 0 | 0 | 0 |
| sub-OAS30421_ses-d2605_run-01_T1w.nii.gz | 0 | 0 | 0 | 0 | 0 | 0 | 0 | 0 | 0 |
| sub-OAS30434_ses-d0054_T1w.nii.gz | 0 | 0 | 0 | 0 | 0 | 0 | 0 | 0 | 0 |
| sub-OAS30436_ses-d0388_run-01_T1w.nii.gz | 0 | II | 0 | 0 | II | 0 | 0 | 0 | 0 |
| sub-OAS30438_ses-d0897_T1w.nii.gz | 0 | II | 0 | 0 | 0 | 0 | 0 | 0 | 0 |
| sub-OAS30440_ses-d0163_T1w.nii.gz | 0 | II | 0 | 0 | II | 0 | 0 | 0 | 0 |

Table S1: Results of the manual review of the 179 $T_1$-weighted images from the OASIS-3 cohort. **Key:** 0 – Success; I – Type I Failure;
II – Type II Failure.

| **Image** | Defacer | Quickshear | Pydeface | MRI_Deface | FSL_deface | Face_Masking | AnonyMI | mri_reface_0.2 | mri_reface_0.3 |
|---|---|---|---|---|---|---|---|---|---|
| sub-OAS30451_ses-d2656_T1w.nii.gz | 0 | 0 | 0 | 0 | II | 0 | 0 | 0 | 0 |
| sub-OAS30452_ses-d4501_T1w.nii.gz | 0 | II | 0 | 0 | 0 | 0 | 0 | 0 | 0 |
| sub-OAS30460_ses-d0043_T1w.nii.gz | 0 | II | 0 | 0 | II | 0 | 0 | 0 | 0 |
| sub-OAS30465_ses-d0152_T1w.nii.gz | 0 | II | 0 | II | 0 | 0 | 0 | 0 | 0 |
| sub-OAS30474_ses-d0069_run-01_T1w.nii.gz | 0 | 0 | 0 | 0 | 0 | 0 | 0 | 0 | 0 |
| sub-OAS30476_ses-d0090_T1w.nii.gz | I | II | 0 | 0 | 0 | 0 | 0 | 0 | 0 |
| sub-OAS30479_ses-d0398_run-01_T1w.nii.gz | 0 | II | 0 | 0 | 0 | 0 | 0 | 0 | 0 |
| sub-OAS30483_ses-d0020_run-01_T1w.nii.gz | 0 | 0 | 0 | 0 | 0 | 0 | 0 | 0 | 0 |
| sub-OAS30486_ses-d1295_T1w.nii.gz | 0 | II | 0 | 0 | 0 | 0 | 0 | 0 | 0 |
| sub-OAS30490_ses-d0412_run-01_T1w.nii.gz | 0 | II | 0 | II | 0 | 0 | 0 | 0 | 0 |
| sub-OAS30494_ses-d4030_T1w.nii.gz | 0 | II | 0 | 0 | 0 | 0 | 0 | 0 | 0 |
| sub-OAS30498_ses-d0132_T1w.nii.gz | 0 | II | 0 | 0 | 0 | 0 | 0 | 0 | 0 |
| sub-OAS30503_ses-d0000_T1w.nii.gz | 0 | II | 0 | 0 | 0 | 0 | 0 | 0 | 0 |
| sub-OAS30507_ses-d2941_T1w.nii.gz | 0 | II | 0 | II | 0 | 0 | 0 | 0 | 0 |
| sub-OAS30510_ses-d1154_T1w.nii.gz | 0 | II | 0 | II | II | 0 | 0 | 0 | 0 |
| sub-OAS30513_ses-d0119_T1w.nii.gz | 0 | 0 | 0 | 0 | 0 | 0 | 0 | 0 | 0 |
| sub-OAS30535_ses-d0139_run-01_T1w.nii.gz | 0 | 0 | 0 | 0 | 0 | 0 | 0 | 0 | 0 |
| sub-OAS30549_ses-d1244_run-01_T1w.nii.gz | I | II | 0 | 0 | 0 | 0 | 0 | 0 | 0 |
| sub-OAS30554_ses-d0118_run-01_T1w.nii.gz | 0 | 0 | 0 | 0 | 0 | 0 | 0 | 0 | 0 |
| sub-OAS30557_ses-d1448_run-01_T1w.nii.gz | I | 0 | 0 | 0 | 0 | 0 | 0 | 0 | 0 |
| sub-OAS30560_ses-d1203_run-01_T1w.nii.gz | 0 | II | 0 | 0 | II | 0 | 0 | 0 | 0 |

Table S1: Results of the manual review of the 179 $T_1$-weighted images from the OASIS-3 cohort. **Key:** 0 – Success; I – Type I Failure;
II – Type II Failure.

| **Image** | Defacer | Quickshear | Pydeface | MRI_Deface | FSL_deface | Face_Masking | AnonyMI | mri_reface_0.2 | mri_reface_0.3 |
|---|---|---|---|---|---|---|---|---|---|
| sub-OAS30567_ses-d0040_T1w.nii.gz | 0 | 0 | 0 | II | 0 | 0 | 0 | 0 | 0 |
| sub-OAS30583_ses-d0085_run-01_T1w.nii.gz | I | II | 0 | II | 0 | 0 | 0 | 0 | 0 |
| sub-OAS30589_ses-d3191_T1w.nii.gz | 0 | 0 | 0 | II | 0 | 0 | 0 | 0 | 0 |
| sub-OAS30596_ses-d2477_T1w.nii.gz | 0 | 0 | 0 | 0 | 0 | 0 | 0 | 0 | 0 |
| sub-OAS30616_ses-d0199_T1w.nii.gz | 0 | 0 | 0 | 0 | 0 | 0 | 0 | 0 | 0 |
| sub-OAS30617_ses-d0073_T1w.nii.gz | 0 | II | 0 | 0 | II | 0 | 0 | 0 | 0 |
| sub-OAS30623_ses-d0054_run-01_T1w.nii.gz | 0 | 0 | 0 | 0 | 0 | 0 | 0 | 0 | 0 |
| sub-OAS30636_ses-d0056_T1w.nii.gz | 0 | II | 0 | 0 | II | 0 | 0 | 0 | 0 |
| sub-OAS30637_ses-d0079_T1w.nii.gz | 0 | II | 0 | II | 0 | 0 | 0 | 0 | 0 |
| sub-OAS30649_ses-d0098_run-01_T1w.nii.gz | 0 | 0 | 0 | 0 | 0 | 0 | 0 | 0 | 0 |
| sub-OAS30652_ses-d0778_run-01_T1w.nii.gz | 0 | II | 0 | 0 | 0 | 0 | 0 | 0 | 0 |
| sub-OAS30653_ses-d0102_run-01_T1w.nii.gz | 0 | 0 | 0 | 0 | 0 | 0 | 0 | 0 | 0 |
| sub-OAS30656_ses-d2737_T1w.nii.gz | 0 | II | 0 | II | 0 | 0 | 0 | 0 | 0 |
| sub-OAS30657_ses-d0072_run-01_T1w.nii.gz | 0 | 0 | 0 | 0 | 0 | 0 | 0 | 0 | 0 |
| sub-OAS30667_ses-d0888_T1w.nii.gz | 0 | II | 0 | 0 | 0 | 0 | 0 | 0 | 0 |
| sub-OAS30673_ses-d2417_run-01_T1w.nii.gz | 0 | II | 0 | 0 | 0 | 0 | 0 | 0 | 0 |
| sub-OAS30677_ses-d0000_T1w.nii.gz | 0 | II | 0 | 0 | 0 | 0 | 0 | 0 | 0 |
| sub-OAS30681_ses-d0154_T1w.nii.gz | 0 | II | 0 | II | II | 0 | 0 | 0 | 0 |
| sub-OAS30685_ses-d1552_T1w.nii.gz | I | 0 | 0 | 0 | 0 | 0 | 0 | 0 | 0 |
| sub-OAS30699_ses-d1809_run-01_T1w.nii.gz | 0 | II | 0 | 0 | 0 | 0 | 0 | 0 | 0 |
| sub-OAS30706_ses-d0060_T1w.nii.gz | 0 | 0 | 0 | 0 | 0 | 0 | 0 | 0 | 0 |

Table S1: Results of the manual review of the 179 $T_1$-weighted images from the OASIS-3 cohort. **Key:** 0 – Success; I – Type I Failure;
II – Type II Failure.

| **Image** | Defacer | Quickshear | Pydeface | MRI_Deface | FSL_deface | Face_Masking | AnonyMI | mri_reface_0.2 | mri_reface_0.3 |
|---|---|---|---|---|---|---|---|---|---|
| sub-OAS30708_ses-d0071_T1w.nii.gz | 0 | 0 | 0 | 0 | 0 | 0 | 0 | 0 | 0 |
| sub-OAS30716_ses-d0037_T1w.nii.gz | 0 | II | 0 | 0 | 0 | 0 | 0 | 0 | 0 |
| sub-OAS30729_ses-d4384_run-01_T1w.nii.gz | 0 | 0 | 0 | 0 | 0 | 0 | 0 | 0 | 0 |
| sub-OAS30732_ses-d0074_T1w.nii.gz | 0 | II | 0 | 0 | II | 0 | 0 | 0 | 0 |
| sub-OAS30755_ses-d0063_run-01_T1w.nii.gz | 0 | II | 0 | 0 | 0 | 0 | 0 | 0 | 0 |
| sub-OAS30756_ses-d1267_T1w.nii.gz | 0 | II | 0 | II | 0 | 0 | 0 | 0 | 0 |
| sub-OAS30757_ses-d2279_T1w.nii.gz | 0 | II | 0 | II | II | 0 | 0 | 0 | 0 |
| sub-OAS30759_ses-d0063_T1w.nii.gz | 0 | 0 | 0 | 0 | 0 | 0 | 0 | 0 | 0 |
| sub-OAS30760_ses-d3837_run-01_T1w.nii.gz | 0 | II | 0 | 0 | II | 0 | 0 | 0 | 0 |
| sub-OAS30762_ses-d1002_T1w.nii.gz | 0 | II | 0 | II | 0 | 0 | 0 | 0 | 0 |
| sub-OAS30776_ses-d2471_T1w.nii.gz | 0 | II | 0 | 0 | II | 0 | 0 | 0 | 0 |
| sub-OAS30783_ses-d0056_run-01_T1w.nii.gz | I | II | 0 | 0 | II | 0 | 0 | 0 | 0 |
| sub-OAS30800_ses-d0029_run-01_T1w.nii.gz | 0 | II | 0 | 0 | 0 | 0 | 0 | 0 | 0 |
| sub-OAS30805_ses-d5456_run-01_T1w.nii.gz | 0 | II | 0 | 0 | 0 | 0 | 0 | 0 | 0 |
| sub-OAS30812_ses-d0055_T1w.nii.gz | 0 | II | 0 | II | 0 | 0 | 0 | 0 | 0 |
| sub-OAS30821_ses-d0063_T1w.nii.gz | 0 | 0 | 0 | 0 | 0 | 0 | 0 | 0 | 0 |
| sub-OAS30832_ses-d2382_T1w.nii.gz | 0 | 0 | 0 | 0 | 0 | 0 | 0 | 0 | 0 |
| sub-OAS30836_ses-d0094_T1w.nii.gz | 0 | II | 0 | 0 | 0 | 0 | 0 | 0 | 0 |
| sub-OAS30842_ses-d0526_T1w.nii.gz | 0 | II | 0 | 0 | 0 | 0 | 0 | 0 | 0 |
| sub-OAS30849_ses-d0151_T1w.nii.gz | 0 | II | 0 | 0 | II | 0 | 0 | 0 | 0 |
| sub-OAS30857_ses-d0058_run-01_T1w.nii.gz | 0 | II | 0 | 0 | 0 | 0 | 0 | 0 | 0 |

Table S1: Results of the manual review of the 179 $T_1$-weighted images from the OASIS-3 cohort. **Key:** 0 – Success; I – Type I Failure; II – Type II Failure.

| **Image** | Defacer | Quickshear | Pydeface | MRI_Deface | FSL_deface | Face_Masking | AnonyMI | mri_reface_0.2 | mri_reface_0.3 |
|---|---|---|---|---|---|---|---|---|---|
| sub-OAS30864_ses-d0603_T1w.nii.gz | I | II | 0 | 0 | 0 | 0 | 0 | 0 | 0 |
| sub-OAS30865_ses-d0109_run-01_T1w.nii.gz | 0 | 0 | 0 | 0 | 0 | 0 | 0 | 0 | 0 |
| sub-OAS30867_ses-d1398_run-01_T1w.nii.gz | 0 | 0 | 0 | 0 | 0 | 0 | 0 | 0 | 0 |
| sub-OAS30872_ses-d1285_run-01_T1w.nii.gz | 0 | II | 0 | I | 0 | II | 0 | 0 | 0 |
| sub-OAS30882_ses-d0191_T1w.nii.gz | 0 | II | 0 | II | 0 | 0 | 0 | 0 | 0 |
| sub-OAS30886_ses-d0045_run-01_T1w.nii.gz | 0 | 0 | 0 | 0 | 0 | 0 | 0 | 0 | 0 |
| sub-OAS30887_ses-d1407_T1w.nii.gz | 0 | 0 | 0 | II | 0 | 0 | 0 | 0 | 0 |
| sub-OAS30908_ses-d0617_run-01_T1w.nii.gz | 0 | II | 0 | 0 | 0 | 0 | 0 | 0 | 0 |
| sub-OAS30910_ses-d0090_T1w.nii.gz | I | II | 0 | 0 | 0 | 0 | 0 | 0 | 0 |
| sub-OAS30915_ses-d0085_T1w.nii.gz | 0 | II | 0 | 0 | 0 | 0 | 0 | 0 | 0 |
| sub-OAS30925_ses-d0196_run-01_T1w.nii.gz | 0 | II | 0 | 0 | 0 | 0 | 0 | 0 | 0 |
| sub-OAS30927_ses-d0145_T1w.nii.gz | 0 | 0 | 0 | I | II | 0 | 0 | 0 | 0 |
| sub-OAS30935_ses-d0201_T1w.nii.gz | 0 | II | 0 | 0 | 0 | 0 | 0 | 0 | 0 |
| sub-OAS30949_ses-d1418_run-01_T1w.nii.gz | 0 | II | 0 | 0 | 0 | 0 | 0 | 0 | 0 |
| sub-OAS30951_ses-d3786_T1w.nii.gz | 0 | II | 0 | 0 | 0 | 0 | 0 | 0 | 0 |
| sub-OAS30962_ses-d0007_T1w.nii.gz | I | II | 0 | I | 0 | 0 | 0 | 0 | 0 |
| sub-OAS30964_ses-d1135_run-01_T1w.nii.gz | 0 | 0 | 0 | 0 | 0 | 0 | 0 | 0 | 0 |
| sub-OAS30978_ses-d0059_run-01_T1w.nii.gz | 0 | 0 | 0 | II | II | 0 | 0 | 0 | 0 |
| sub-OAS30986_ses-d2308_T1w.nii.gz | 0 | 0 | 0 | 0 | 0 | 0 | 0 | 0 | 0 |
| sub-OAS30989_ses-d0109_run-01_T1w.nii.gz | 0 | 0 | 0 | 0 | 0 | 0 | 0 | 0 | 0 |
| sub-OAS30993_ses-d0920_T1w.nii.gz | 0 | II | 0 | II | II | 0 | 0 | 0 | 0 |

Table S1: Results of the manual review of the 179 $T_1$-weighted images from the OASIS-3 cohort. **Key:** 0 – Success; I – Type I Failure;
II – Type II Failure.

| **Image** | Defacer | Quickshear | Pydeface | MRI_Deface | FSL_deface | Face_Masking | AnonyMI | mri_reface_0.2 | mri_reface_0.3 |
|---|---|---|---|---|---|---|---|---|---|
| sub-OAS30997_ses-d0058_T1w.nii.gz | 0 | II | 0 | 0 | II | 0 | 0 | 0 | 0 |
| sub-OAS31004_ses-d0000_T1w.nii.gz | I | 0 | 0 | 0 | II | 0 | 0 | 0 | 0 |
| sub-OAS31006_ses-d1106_T1w.nii.gz | 0 | 0 | 0 | 0 | II | 0 | 0 | 0 | 0 |
| sub-OAS31007_ses-d0304_run-01_T1w.nii.gz | 0 | II | 0 | 0 | II | 0 | 0 | 0 | 0 |
| sub-OAS31009_ses-d0512_run-01_T1w.nii.gz | 0 | 0 | 0 | 0 | 0 | II | 0 | 0 | 0 |
| sub-OAS31029_ses-d2448_T1w.nii.gz | 0 | II | 0 | 0 | 0 | 0 | 0 | 0 | 0 |
| sub-OAS31035_ses-d5659_run-01_T1w.nii.gz | 0 | II | 0 | 0 | 0 | 0 | 0 | 0 | 0 |
| sub-OAS31038_ses-d4037_run-01_T1w.nii.gz | 0 | 0 | 0 | 0 | 0 | 0 | 0 | 0 | 0 |
| sub-OAS31051_ses-d0365_run-01_T1w.nii.gz | 0 | 0 | 0 | 0 | 0 | 0 | 0 | 0 | 0 |
| sub-OAS31054_ses-d2787_T1w.nii.gz | 0 | 0 | 0 | II | II | 0 | 0 | 0 | 0 |
| sub-OAS31063_ses-d0146_run-01_T1w.nii.gz | 0 | II | 0 | 0 | 0 | 0 | 0 | 0 | 0 |
| sub-OAS31069_ses-d0025_run-01_T1w.nii.gz | 0 | 0 | 0 | 0 | 0 | 0 | 0 | 0 | 0 |
| sub-OAS31078_ses-d0027_run-01_T1w.nii.gz | 0 | II | 0 | 0 | 0 | 0 | 0 | 0 | 0 |
| sub-OAS31085_ses-d0112_run-01_T1w.nii.gz | 0 | 0 | 0 | 0 | 0 | 0 | 0 | 0 | 0 |
| sub-OAS31086_ses-d0065_T1w.nii.gz | 0 | II | 0 | I | 0 | 0 | 0 | 0 | 0 |
| sub-OAS31087_ses-d1461_T1w.nii.gz | 0 | 0 | 0 | 0 | 0 | 0 | 0 | 0 | 0 |
| sub-OAS31093_ses-d0063_run-01_T1w.nii.gz | 0 | 0 | 0 | 0 | 0 | 0 | 0 | 0 | 0 |
| sub-OAS31094_ses-d0103_run-01_T1w.nii.gz | 0 | 0 | 0 | 0 | 0 | 0 | 0 | 0 | 0 |
| sub-OAS31097_ses-d2763_run-01_T1w.nii.gz | 0 | 0 | 0 | 0 | 0 | 0 | 0 | 0 | 0 |
| sub-OAS31105_ses-d1017_run-01_T1w.nii.gz | 0 | 0 | 0 | 0 | 0 | 0 | 0 | 0 | 0 |
| sub-OAS31106_ses-d0065_run-01_T1w.nii.gz | 0 | 0 | 0 | 0 | 0 | 0 | 0 | 0 | 0 |

Table S1: Results of the manual review of the 179 $T_1$-weighted images from the OASIS-3 cohort. **Key:** 0 – Success; I – Type I Failure;
II – Type II Failure.

| **Image** | Defacer | Quickshear | Pydeface | MRI_Deface | FSL_deface | Face_Masking | AnonyMI | mri_reface_0.2 | mri_reface_0.3 |
|---|---|---|---|---|---|---|---|---|---|
| sub-OAS31107_ses-d0912_T1w.nii.gz | 0 | II | 0 | 0 | 0 | 0 | 0 | 0 | 0 |
| sub-OAS31112_ses-d0276_run-01_T1w.nii.gz | 0 | 0 | 0 | 0 | 0 | 0 | 0 | 0 | 0 |
| sub-OAS31114_ses-d1442_run-01_T1w.nii.gz | 0 | 0 | 0 | 0 | 0 | 0 | 0 | 0 | 0 |
| sub-OAS31115_ses-d0466_T1w.nii.gz | 0 | 0 | 0 | II | II | 0 | 0 | 0 | 0 |
| sub-OAS31117_ses-d0073_T1w.nii.gz | 0 | II | 0 | II | II | 0 | 0 | 0 | 0 |
| sub-OAS31123_ses-d0078_T1w.nii.gz | 0 | 0 | 0 | 0 | 0 | 0 | 0 | 0 | 0 |
| sub-OAS31126_ses-d1156_run-01_T1w.nii.gz | 0 | II | 0 | 0 | 0 | 0 | 0 | 0 | 0 |
| sub-OAS31132_ses-d3576_run-01_T1w.nii.gz | 0 | II | 0 | 0 | II | 0 | 0 | 0 | 0 |
| sub-OAS31137_ses-d3443_T1w.nii.gz | 0 | II | 0 | 0 | 0 | 0 | 0 | 0 | 0 |
| sub-OAS31141_ses-d0106_run-01_T1w.nii.gz | 0 | II | 0 | 0 | 0 | 0 | 0 | 0 | 0 |
| sub-OAS31147_ses-d0147_T1w.nii.gz | I | II | 0 | II | II | 0 | 0 | 0 | 0 |

Table S2: Results of the manual review of the 21 $T_1$-weighted images from the Kirby-21 dataset. **Key:** 0 – Success; I – Type I Failure;
II – Type II Failure.

| **Image** | Defacer | Quickshear | Pydeface | MRI_Deface | FSL_deface | Face_Masking | AnonyMI | mri_reface_0.2 | mri_reface_0.3 |
|---|---|---|---|---|---|---|---|---|---|
| KKI2009-01-MPRAGE | 0 | 0 | 0 | 0 | 0 | 0 | 0 | 0 | 0 |
| KKI2009-02-MPRAGE | 0 | 0 | 0 | 0 | 0 | 0 | 0 | 0 | 0 |
| KKI2009-03-MPRAGE | 0 | 0 | 0 | 0 | 0 | 0 | 0 | 0 | 0 |
| KKI2009-04-MPRAGE | 0 | 0 | 0 | 0 | 0 | 0 | 0 | 0 | 0 |
| KKI2009-05-MPRAGE | 0 | 0 | 0 | 0 | 0 | 0 | 0 | 0 | 0 |
| KKI2009-06-MPRAGE | 0 | 0 | 0 | 0 | 0 | 0 | 0 | 0 | 0 |
| KKI2009-07-MPRAGE | 0 | 0 | 0 | 0 | 0 | 0 | 0 | 0 | 0 |
| KKI2009-08-MPRAGE | 0 | 0 | 0 | 0 | 0 | 0 | 0 | 0 | 0 |
| KKI2009-09-MPRAGE | 0 | 0 | 0 | 0 | 0 | 0 | 0 | 0 | 0 |
| KKI2009-10-MPRAGE | 0 | 0 | 0 | 0 | 0 | 0 | 0 | 0 | 0 |
| KKI2009-12-MPRAGE | 0 | 0 | 0 | 0 | 0 | 0 | 0 | 0 | 0 |
| KKI2009-13-MPRAGE | 0 | 0 | 0 | 0 | 0 | 0 | 0 | 0 | 0 |
| KKI2009-14-MPRAGE | 0 | 0 | 0 | 0 | 0 | 0 | 0 | 0 | 0 |
| KKI2009-15-MPRAGE | 0 | 0 | 0 | 0 | 0 | 0 | 0 | 0 | 0 |
| KKI2009-16-MPRAGE | 0 | 0 | 0 | 0 | 0 | 0 | 0 | 0 | 0 |
| KKI2009-18-MPRAGE | 0 | 0 | 0 | 0 | 0 | 0 | 0 | 0 | 0 |
| KKI2009-23-MPRAGE | 0 | 0 | 0 | 0 | 0 | 0 | 0 | 0 | 0 |
| KKI2009-28-MPRAGE | 0 | 0 | 0 | 0 | 0 | 0 | 0 | 0 | 0 |
| KKI2009-30-MPRAGE | 0 | 0 | 0 | 0 | 0 | 0 | 0 | 0 | 0 |
| KKI2009-32-MPRAGE | 0 | 0 | 0 | 0 | 0 | 0 | 0 | 0 | 0 |
| KKI2009-39-MPRAGE | 0 | II | 0 | 0 | 0 | 0 | 0 | 0 | 0 |

### List of Tables

S1 Results of the manual review of the 179 $T_1$-weighted images from the OASIS-3 cohort. **Key:** 0 – Success; I – Type I Failure; II – Type II Failure. 13

S1 Results of the manual review of the 179 $T_1$-weighted images from the OASIS-3 cohort. **Key:** 0 – Success; I – Type I Failure; II – Type II Failure. 14

S1 Results of the manual review of the 179 $T_1$-weighted images from the OASIS-3 cohort. **Key:** 0 – Success; I – Type I Failure; II – Type II Failure. 15

S1 Results of the manual review of the 179 $T_1$-weighted images from the OASIS-3 cohort. **Key:** 0 – Success; I – Type I Failure; II – Type II Failure. 16

S1 Results of the manual review of the 179 $T_1$-weighted images from the OASIS-3 cohort. **Key:** 0 – Success; I – Type I Failure; II – Type II Failure. 17

S1 Results of the manual review of the 179 $T_1$-weighted images from the OASIS-3 cohort. **Key:** 0 – Success; I – Type I Failure; II – Type II Failure. 18

S1 Results of the manual review of the 179 $T_1$-weighted images from the OASIS-3 cohort. **Key:** 0 – Success; I – Type I Failure; II – Type II Failure. 19

S1 Results of the manual review of the 179 $T_1$-weighted images from the OASIS-3 cohort. **Key:** 0 – Success; I – Type I Failure; II – Type II Failure. 20
S1 Results of the manual review of the 179 $T_1$-weighted images from the OASIS-3 cohort. **Key:** 0 – Success; I – Type I Failure; II – Type II Failure. 21
S2 Results of the manual review of the 21 $T_1$-weighted images from the Kirby-21 dataset. **Key:** 0 – Success; I – Type I Failure; II – Type II Failure. . . 22
